# Genomic Epidemiology of Respiratory Syncytial Virus in a New England Hospital System, 2024

**DOI:** 10.1101/2025.02.21.25322621

**Authors:** Emily LaVerriere, Sasha Behar, Cole Sher-Jan, Yan Mei Liang, Manish Sagar, John H. Connor

**Author notes:** These authors contributed equally to this work (Emily LaVerriere, Sasha Behar). Corresponding author: John H. Connor. 620 Albany St., Boston, MA 02118.

## Abstract

Respiratory syncytial virus (RSV) is one of the main seasonal respiratory pathogens in the United States and causes up to 240,000 hospitalizations per year among children under five and adults over 60. RSV is classified into two subtypes, RSV-A and RSV-B. Although several RSV vaccines and a preventative monoclonal antibody, nirsevimab, were recently approved by the FDA, vaccination and prevention measures remain low even among age groups at higher risk for hospitalization. To better understand the epidemiology of RSV, we analyzed RSV-positive nasopharyngeal swabs from Boston Medical Center and its satellite clinics from January to June 2024 using amplicon-based whole-genome sequencing. Of the 59 samples collected, 19 were from children under five years of age, and 17 were from adults over the age of 60. Fifty-four samples sequenced successfully, with over 90% of the genome at a minimum of 20-fold coverage.

We found that over 80% of the samples were RSV-B; 48 RSV-B samples and 6 RSV-A samples. This represents a major switch from 2022, when Boston RSV samples were ~90% RSV-A. 45 of 48 RSV-B samples mapped into a single clade (B.D.E.1). These samples do not cluster to a single source within the B.D.E.1 clade, suggesting that the predominance of RSV-B is multifactorial, not the selective expansion of a single variant. We also found examples of identical/highly related genomes among our samples, suggesting clustered transmission. One infant had documented nirsevimab therapy forty days prior to RSV isolation. None of the adults had documented RSV vaccination, and mutations associated with nirsevimab resistance or vaccine escape were not observed. Our work highlights the importance of genomic surveillance for respiratory pathogens as a means to monitor transmission dynamics, such as the unexpected switch from RSV-A to RSV-B subtype dominance, to identify examples of superspreading events, and to understand the epidemiological changes that may be associated with nirsevimab and RSV vaccines.

## Introduction

Respiratory syncytial virus (RSV) is an ongoing threat to human health. An estimated 3.2 million infants and children under age five worldwide are hospitalized due to RSV infection each year (80,000 per year in the United States).^1^ Adults over age 65, or those of any age with certain chronic medical conditions, have an elevated risk of severe disease and hospitalization following RSV infection. Estimates from a recent meta-analysis suggest over 150,000 adults over age 65 are hospitalized due to RSV infection each year in the United States.^2^ RSV transmission is generally seasonal, peaking over the winter in temperate regions. The two subtypes of RSV (RSV-A and RSV-B) are most often observed co-circulating during transmission seasons, and whichever subtype dominates in a season has not been associated with temporal variation.^3^

As current clinical tests for RSV infection do not differentiate between the subtypes, whole-genome sequencing has been employed for epidemiological surveillance. Amplicon-based whole genome sequencing is well-suited to amplify target viral RNA within nasal samples containing competing RNA. Multiple tiled amplicon designs have been developed and used for RSV genomic surveillance in recent years.^4-10^

Genomic analysis also allows for investigation of whether circulating RSV is accumulating resistance to therapeutics or vaccines. Three RSV vaccines were recently introduced to the United States general public, for all adults ≥ 75, adults between ages 60-74 with increased risk of severe RSV, and pregnant individuals between weeks 32-36 of pregnancy.^11^ Uptake for vaccination in these three groups in July 2024 was estimated at 30.8% (95% CI: 30.1-31.6), 20.7% (95% CI: 20.2-23.2), and 7.7% (denominator: 12,586), respectively.^12^ Nirsevimab, a monoclonal antibody, was recently recommended in the United States for RSV prophylaxis for infants under age one.^13^ The F (fusion) gene within RSV is the basis of all three current vaccines and contains the binding site for nirsevimab. Although much of the F gene sequence has historically been well conserved^14^, RSV whole-genome sequencing allows us to uncover new possible mutations and enables the detect changes that may occur in response to vaccination and monoclonal antibodies. Recently, a portion of RSV-B breakthrough infections contained mutations associated with nirsevimab resistance^15^, highlighting the need for ongoing surveillance for resistance mutations.

We were interested in understanding RSV circulation in the greater Boston area during the 2024 transmission season. The most recent genomic surveillance for RSV in this area is from 2022, when 90% of sequenced genomes were part of the RSV-A sublineage.^16^ To better understand the currently circulating RSV and how it compares to the 2022 genomes, we collected samples from individuals who tested positive for RSV at Boston Medical Center, a safety-net hospital, and its satellite clinics. Using amplicon-based whole genome sequencing of both RSV-A and RSV-B, we analyzed the patterns of genetic diversity and signals of transmission.

## Methods

### Samples

All nasopharyngeal swabs positive for RSV were identified from the electronic medical record and requested from the BMC clinical microbiology laboratory. Some requested leftover clinical samples could not be obtained because they were either already discarded, missing, or had inadequate quantity. Leftover clinical samples were stored at −80 °C in 300 uL of viral transport medium. All sample and data collection was approved by the Boston University institutional review board (H-42887).

### Sample processing

Each sample was batch-processed through a 96-well plate, beginning with RNA extraction following the Zymo Quick-RNA Viral 96 kit (no DNase I treatment) (catalog: R1040). Prior to the final elution of RNA into nuclease-free water, samples were subjected to a dry spin (no solution added). cDNA was then generated from the resulting RNA using the New England Biolabs LunaScript® RT SuperMix Kit (catalog: E3010). Tiling amplicons ranging in 100-400 base pairs in length were produced using RSV-A and RSV-B specific amplicon primers designed by the Lauring Lab at University of Michigan, Ann Arbor^17^ resulting in four distinct primer pools. Each pool was run on a 1.5% agarose gel to confirm the presence of appropriate amplicon fragments. Successful samples were pooled and underwent Illumina Nextera XT DNA Library Preparation (catalog: FC-131-1096) for next-generation sequencing using an Illumina NextSeq 2000.

### Sequencing data processing

For each sample, we aligned reads to both the RSV-A and RSV-B reference genomes (EPI_ISL_412866 and EPI_ISL_1653999, respectively) using minimap2^18^. We trimmed primer-binding sites and assessed depth of the aligned reads using samtools^19^. We called consensus sequences using samtools, requiring a minimum read depth of 20 and a call fraction above 50%. We called variants using LoFreq^20^, again requiring a minimum read depth of 20. We annotated variants and gene positions with a custom R script, using the tidyverse^21^ and Biostrings^22^ packages. Variants used in all analyses were at consensus within a given sample, with the exception of the SNVs from genome BMC_RSV_145, included in Supplemental Table 1, which included subconsensus SNVs above 10% within-sample frequency.

### Data analysis

We used Nextclade^23^ to generate coverage statistics and assign G-protein^24^ and full-genome clades to each sample, following the standardized phylogenetic classification defined recently^25^. We performed all multiple sequence alignment using Clustal Omega.^26^ We generated maximum-likelihood trees using ModelFinder^27^ within IQTree v2.3.6^28^, as well as time-resolved trees using the least square dating method^29^ within IQTree. We used a custom R script to make pairwise comparisons between genomes, adapted from previous scripts^30^, using R 4.4.1^26^.

We compared consensus sequences from the newly sequenced genomes to genomes from GISAID. On November 8, 2024, we filtered all GISAID RSV genomes available to those flagged as both “complete” and “high coverage.” We manually reviewed alignments of the RSV-A and RSV-B genomes and removed 13 RSV-B genomes from the analysis, resulting in final comparator datasets of 600 RSV-A and 796 RSV-B genomes. The full list of genomes with GISAID IDs is in Supplemental Table 2. We also included 7 RSV-B genomes from the surveillance of the Boston 2022 RSV surge^16^; the GenBank IDs for these genomes are also in Supplemental Table 2.

We plotted all data in R 4.4.1^20^, with the tidyverse^31^, ggprism^32^, and ggtree suite of packages.^33–37^ We arranged final figures in Adobe Illustrator.

## Results

From January 18, 2024 to June 18, 2024, there were 108 nasal swabs with confirmed RSV using the comprehensive respiratory panel polymerase chain reaction (Biofire Diagnostics), respiratory mini panel PCR (BioMerioux) or Cobas-liat (Roche). All nasal swabs with detected RSV were requested from the BMC clinical laboratory. We received 59 of the clinical leftover swabs. The remaining swabs were not received because they had already been discarded or were not available for other reasons. The demographics of the individuals with sequenced samples was different from those the individuals with unavailable samples (Table 1). The average patient age of sequenced samples was 37 (standard deviation 29), while the average patient age of all RSV-positive samples was 28 (standard deviation 30) (Figure 1A). Temporally, the majority of samples were collected within the conventional RSV season (November-February), peaking in January (Figure 1B).

**Table 1.** Demographics of RSV-positive samples.

| Variable | Level | No. Sequenced | No. Not sequenced | No. Total |
| --- | --- | --- | --- | --- |
| Age (years) | Mean (SD) | 37 (29) | 18 (29) | 28 (30) |
|  | Median | 42 | 1 | 9 |
|  | 0-2 | 17 | 28 | 45 |
|  | 60-74 | 13 | 2 | 15 |
|  | >= 75 | 4 | 4 | 8 |
| Sex | Female | 33 | 23 | 56 |
|  | Male | 26 | 26 | 52 |
| Collection Month (in 2024) | January | 26 | 13 | 39 |
|  | February | 22 | 12 | 34 |
|  | March | 3 | 18 | 21 |
|  | April | 5 | 6 | 11 |
|  | May | 2 | 0 | 2 |
|  | June | 1 | 0 | 1 |

Demographic information for all samples identified for inclusion in this study. The “No. Sequenced” column refers to the 59 samples which were processed for whole-genome sequencing. The “No. Not Sequenced” column refers to the 49 samples which were identified as RSV-positive from the electronic medical record but could not be obtained for sequencing. The “No. Total” column refers to all 108 of these samples together.

**Figure 1.**
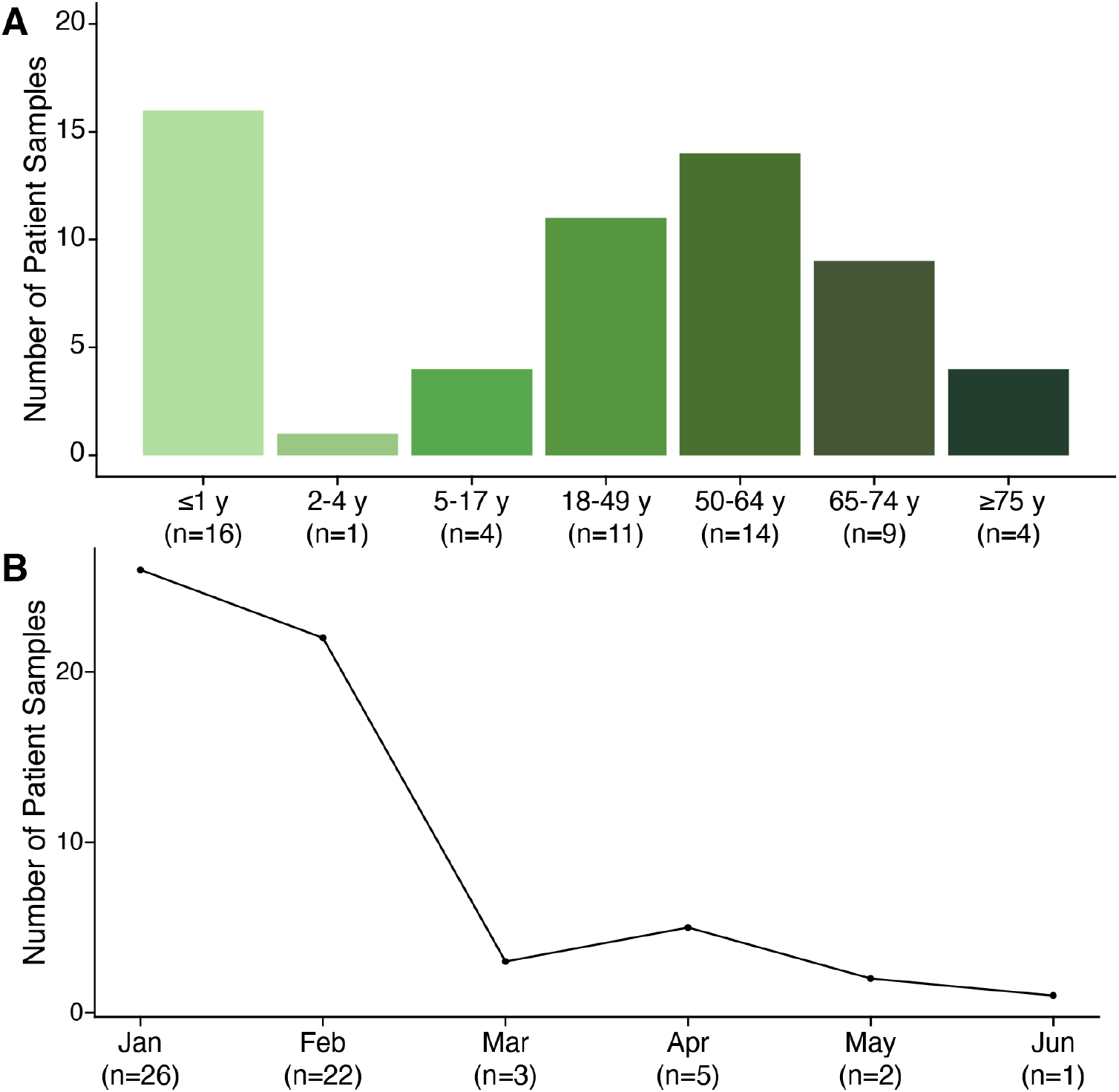
Patient ages and collection dates of RSV samples received from Boston Medical Center in 2024. A) Number of samples received from Boston Medical Center, stratified by patient age group. B) Number of samples received per month (January - June 2024, total n=59).

We used amplicon-based Illumina sequencing to selectively amplify RSV genomes (Figure 2A). We extracted total RNA from the viral transport medium and amplified each sample separately with tiled amplicon primers specific for RSV-A or RSV-B17. We then identified samples with robust amplification for an individual subtype (A or B) for next-generation library preparation and sequencing. We aligned all processed sequencing data to both RSV-A and RSV-B reference genomes. Samples that amplified with RSV-A primers aligned well to the RSV-A genome (median read-depth >1,000) and did not align well to the RSV-B genome (Figure 2B); the reverse was true for samples that amplified with RSV-B primers. We examined coverage across each full genome, as well as across the F and G genes individually (Figure 2C). All samples showed effective amplification and proceeded through library construction (Figure 2D). One sample did not generate usable sequencing data. Of the 58 successfully sequenced genomes, 54 had a minimum of 20x genome coverage across >90% of the full genome and of each the F and G genes. Of these 54 genomes, six were subtype RSV-A and 48 were subtype RSV-B. The RSV-A genomes originated entirely from children ages six and under (Supplemental Figure 1A), while the RSV-B samples originated from individuals of all ages (Supplemental Figure 1B). Both subtypes were detected throughout the sample collection period (Supplemental Figure 1C).

**Figure 2.**
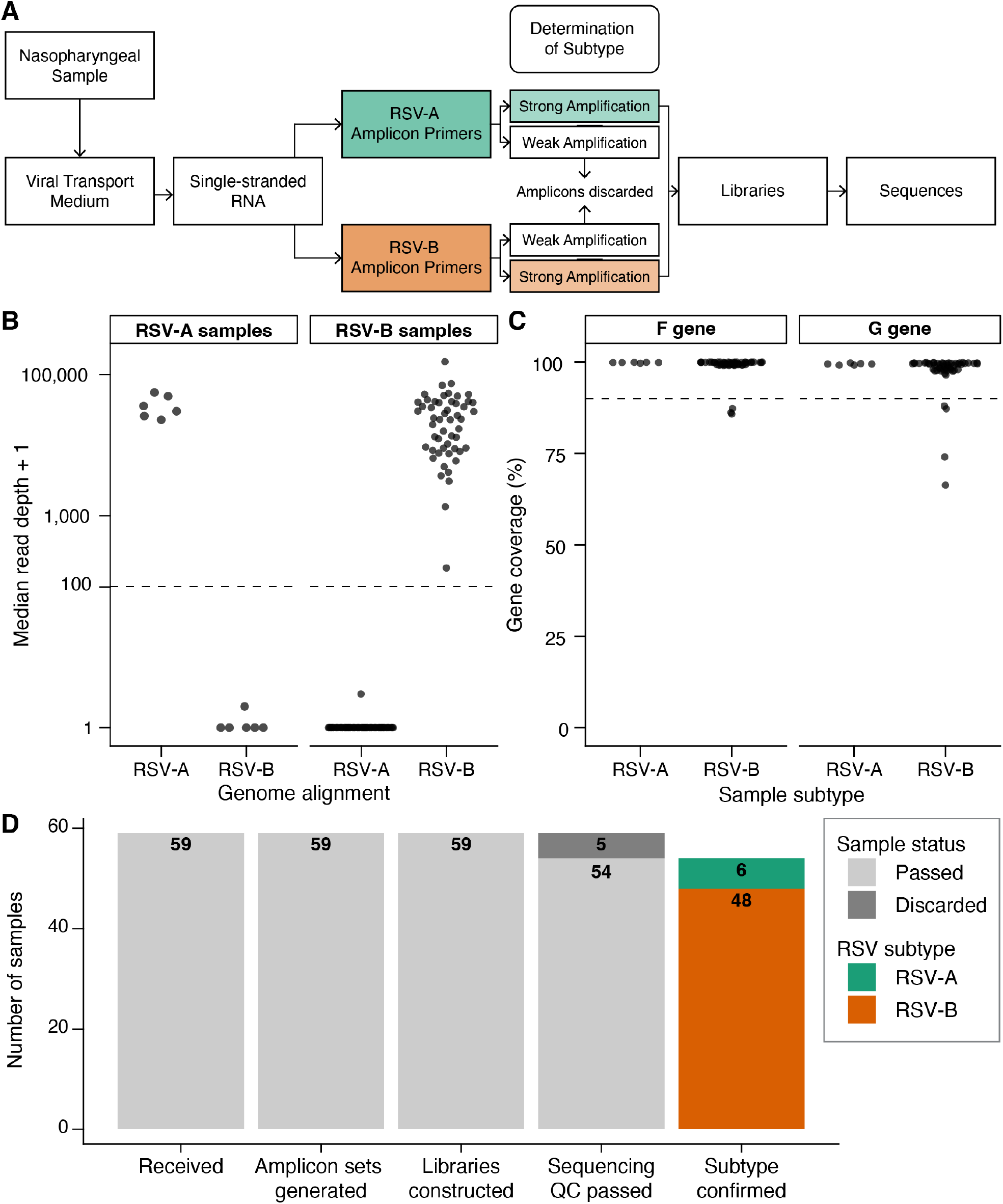
Genomic data generation and quality metrics of sequenced samples. A) The sample processing pipeline from nasopharyngeal sample input to whole-genome sequence data output. B) We aligned processed sequence data from each sample to both RSV-A and RSV-B reference genomes. “RSV-A Samples” describe samples amplified with the RSV-A primer sets, and “RSV-B” Samples” describe samples amplified with the RSV-B primer sets. The y-axis shows read depth + 1, to aid in plotting on a log scale. The dashed line is at 100 read-pairs. C) The y-axis represents the percent of nucleotides within the F or G gene that had > 20x read depth for a given sample. The dashed line at 90% represents the threshold used to retain genomes for further analysis. D) Sample success tracker for the 59 original RSV samples received. The y-axis represents the number of samples and their result at each step of the process. The x-axis shows samples as they proceeded through aliquoting, amplification, library construction, initial sequencing QC, and subtype confirmation (through alignment to reference genomes, as shown in panel B).

We built maximum-likelihood phylogenetic trees for the RSV-A and RSV-B genomes and all high coverage genomes available from GISAID, the global data science initiative^38^, as of November 8, 2024. We also assigned G-protein genotypes^24^ and whole-genome clades^25^ to genomes in the trees using Nextclade^23^. The RSV-B genomes were genetically similar to each other (Figure 3A); they all had the GB5.0.5a G-protein genotype, and nearly all were in the B.D.E.1 clade (45/48). These B.D.E.1 genomes did not form a single cluster; instead, these genomes were representative of much of the global diversity within the B.D.E.1 clade.

**Figure 3.**
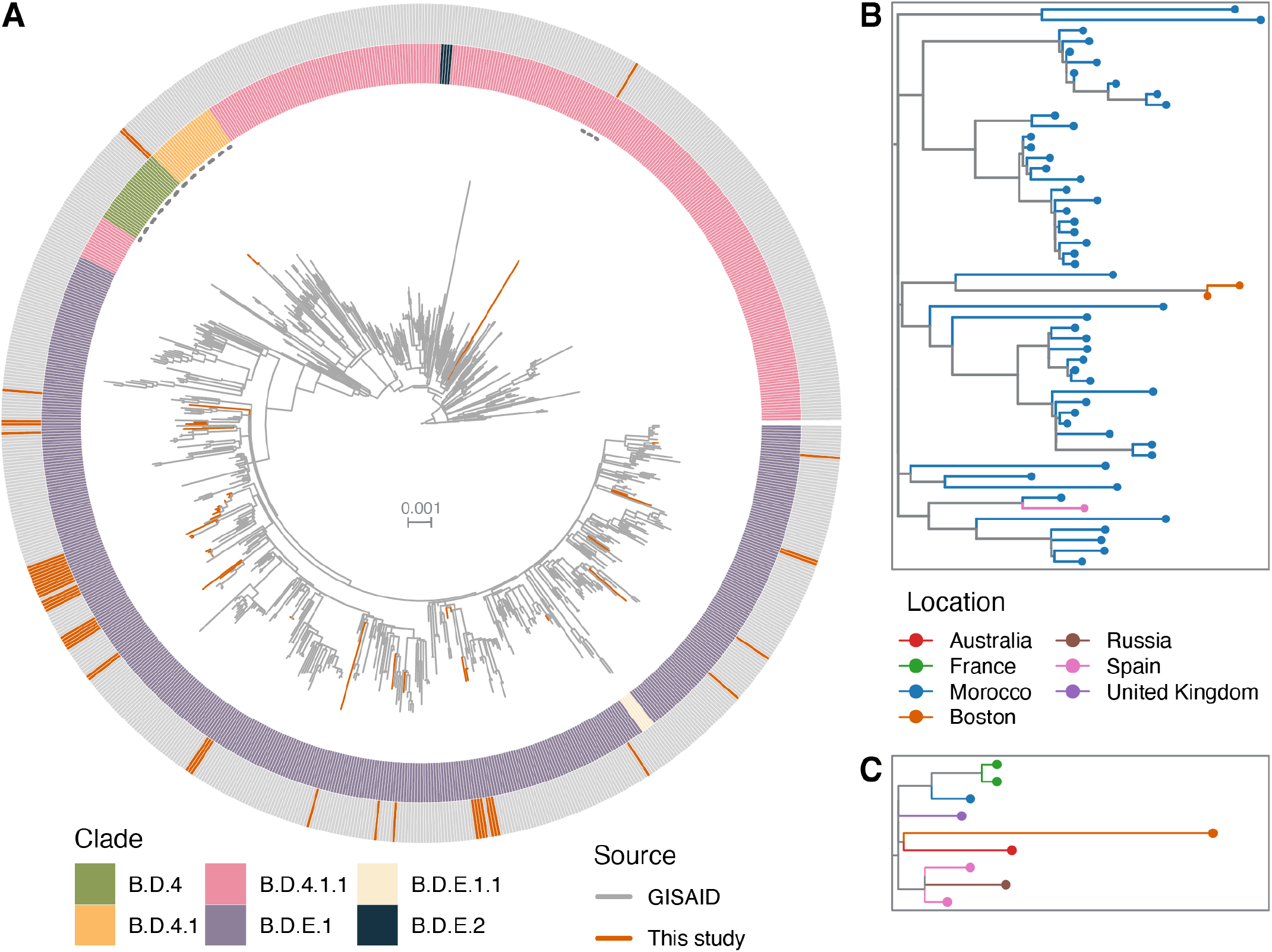
RSV-B genomes from this study compared to global RSV-B genomes. A) Maximum likelihood phylogenetic tree of all RSV-B genomes sequenced in this study (orange) and all high coverage, complete RSV-B genomes available in GISAID as of November 8, 2024 (n=796). The inner ring is colored by each genome’s clade (assigned by Nextclade), and the outer ring is colored by genome source (GISAID in grey, genomes from this study in orange). The dashed lines on the inside of the clade ring denote the regions of the tree that are expanded in panels B and C. B) Expansion of the B.D.4 genomes within the larger tree in panel A. All genomes are colored by location, and the 2 genomes labeled “Boston” and colored in orange are from this study. C) Expansion of a portion of the B.D.4.1.1 genomes within the larger tree in panel A. All genomes are colored by location; the genome labeled “Boston” and colored in orange is from this study.

The non-B.D.E.1 genomes were split between clades B.D.4 (n=2) and B.D.4.1.1 (n=1). When comparing these sequences to other sequences within the GISAID database, we found that the two B.D.4 genomes are most similar to each other, and they fell within a cluster of genomes almost entirely from Morocco (Figure 3B). The single BMC genome within the B.D.4.1.1 clade (Figure 3C) was most similar to a single genome from Australia. The other samples closest to these B.D.4.1.1 genomes were from disparate geographies, suggesting that this genome is from an undersampled area of the RSV-B phylogeny.

To look for potential signs of transmission links, we examined the BMC genomes within the B.D.E.1 clade and their similarities to each other (Figure 4A). We performed pairwise comparisons between genomes in this clade. We found that many of these genomes are unique, but others formed small clusters of highly related genomes (Figure 4B). The first cluster consisted of two samples originating from the same person, taken one day apart, resulted in identical genomes (BMC_RSV_11 and BMC_RSV_20). Another highly related cluster of genomes were from samples taken within a two-week period of time in late January (BMC_RSV_48, BMC_RSV_26, BMC_RSV_50, BMC_RSV_09), suggesting potential transmission links.

**Figure 4.**
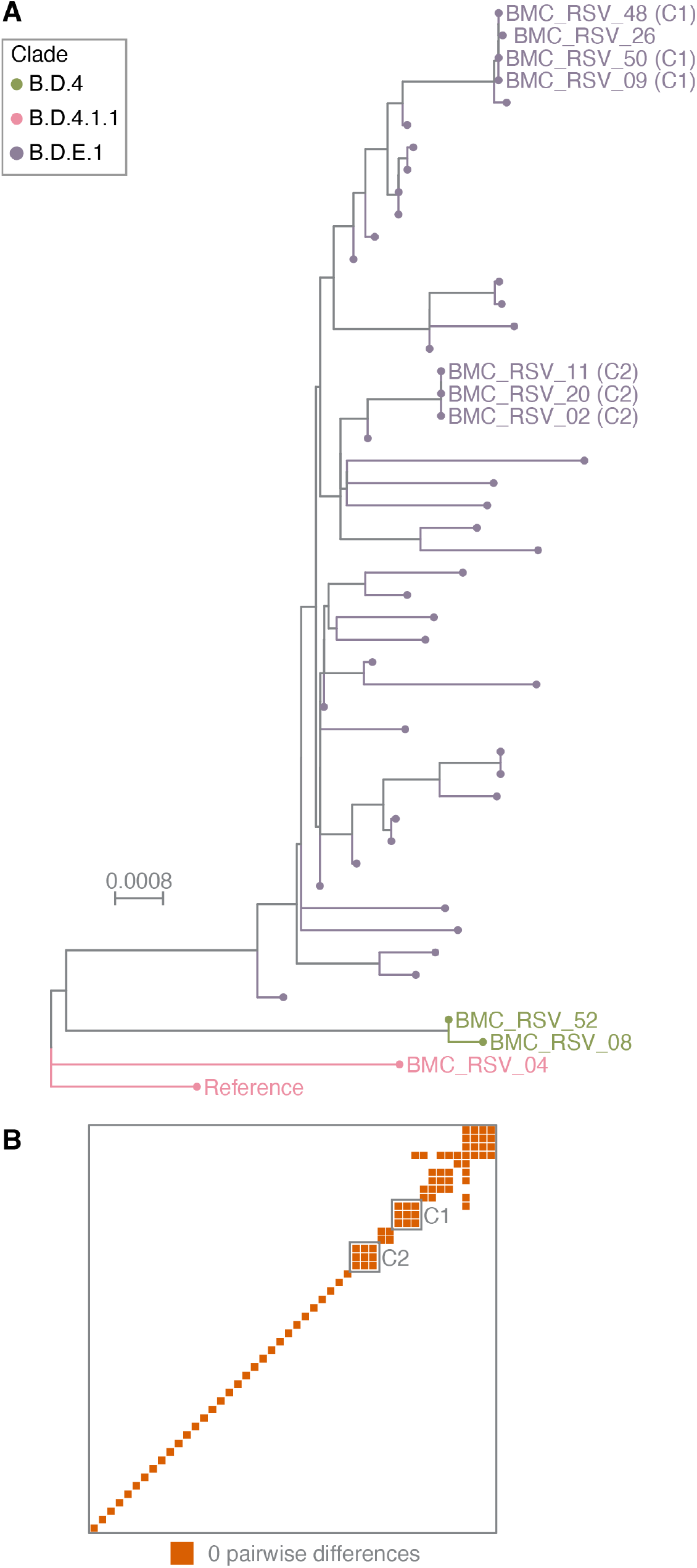
Examination of RSV-B genomes from this study alone. A) Maximum likelihood phylogenetic tree of all RSV-B genomes sequenced in this study, aligned to reference genome (EPI_ISL_1653999, labeled as “Reference” in the figure). Genomes are colored by clade, as assigned by Nextclade. Genomes labeled by sample ID are specifically mentioned in the text. C1 and C2 denote genomes that are part of clusters 1 and 2, respectively. B) Pairwise comparisons of consensus sequences of all RSV-B genomes sequenced in this study. Each genome is represented by a row and column, and each tile is colored by the number of differences between the genomes intersecting at it. Only genome pairs with 0 differences between them are colored (in orange). Grey boxes denote the comparisons between genomes in cluster 1 (C1) and cluster 2 (C2), matching those indicated in panel A.

The RSV-A samples from BMC were a minority of sequenced samples (n=6). They were evenly distributed across the collection period, and the genomes were more genetically diverse than the BMC RSV-B genomes (Supplemental Figure 2). While they all shared the GA2.3.5 G-protein genotype, the six genomes were from five distinct clades (A.D.1, A.D.5.1, A.D.1.4, A.D.1.5, and A.D.1.6). This indicated that there were multiple introductions of RSV-A to the BMC community, but no onward transmission was detected.

All three of the RSV vaccines currently available in the United States contain protein or mRNA of the prefusion form of the F gene. Nirsevimab, a monoclonal antibody used as prophylaxis for infants before the winter transmission season, targets antigenic regions of the F protein. With these potential selection pressures in mind, we examined all non-synonymous single-nucleotide variants (SNVs) within the F gene in both the BMC genomes and all 2024 genomes from GISAID (Supplemental Table 1). We identified 15 SNVs in the BMC samples, seven of which were present in more than one genome. Three SNVs (S190N, S211N, S389P) were present in over 90% of both the BMC and GISAID samples, representing almost all of the B.D.E.1 genomes in each sample set. One individual in the BMC cohort (BMC_RSV_14) received prophylactic nirsevimab 40 days prior to their RSV-positive sample. We examined sub-consensus SNVs within the genome isolated from this sample (Supplemental Table 1). This genome contained three sub-consensus non-synonymous SNVs within the F-gene (L467F, A529V, T558A); however, none of these have been previously associated with nirsevimab resistance.

## Discussion

In this study, we report on RSV genomic diversity in the greater Boston area from January-June 2024. We found an unexpected shift in subtype dominance in this set of genomes compared to what was previously seen in the area in 2022 and compared to other recent RSV surveillance elsewhere in the US. The majority of genomes sequenced in this study were RSV-B (48/54), while the most recent previous RSV genomes from the Boston area were primarily RSV-A (70/77 in 2022)^16^.

Recent surveillance in Arizona^39^, Ontario^8^, and France^40^ reported that RSV-A genomes predominate in their sequencing studies. This is analogous to the 2022 RSV surge described in Boston^16^. Surveillance in Minnesota from July 2023-February 2024^41^, overlapping partially with our sampling period, reported nearly equal numbers of RSV-A and RSV-B genomes, the highest recent proportion of RSV-B genomes in the United States reported before this study. At this time, it is unclear whether the predominance of RSV-B in our study presages a convergence to RSV-B nationwide, or if this predominance is a more geographically restricted phenomenon.

Within the RSV-B genomes sequenced in this study, we found clade representation similar to other recent studies ^8,10,17,39–42^. Nearly all of the Boston RSV-B genomes were within clade B.D.E.1. The B.D.E.1 clade also contains over half of the high coverage, complete genomes available on GISAID as of November 8, 2024. Interestingly, of the seven RSV-B genomes sequenced in the previous Boston dataset^16^, six are in a cluster closely related to a cluster of 14 of the 2024 genomes sequenced in this study (Supplemental Figure 3). This suggests that the RSV-B dominance in our study was not solely from expansion of virus populations from 2022; instead, multiple RSV-B strains were introduced that have continued to circulate during the 2024 season.

The three RSV-B genomes that did not map into the B.D.E.1 clade had recent common ancestors that suggested an origin outside of the US. This indicates but does not prove that these cases could be travel-related. Importantly, the introduction of these distinct RSV-B clade genomes did not lead to further onward transmission within the BMC community. The predominance of B.D.E.1 in our dataset and the minimal clade dispersion is consistent with transmission of RSV-B being driven by local and not imported transmission. The predominance of B.D.E.1 was different from what we and others observe with the RSV-A genome. We identified multiple RSV-A clades in circulation, similar to the diversity of clades present in recent RSV-A surveillance in North America^8,41^ and globally^10^.

In addition to these patterns of genetic diversity, we examined polymorphisms within the F gene to see if there was evidence of purifying selection for mutations that would provide resistance to FDA-approved monoclonal antibodies or RSV vaccines. However, the consistency in common nonsynonymous SNVs in the samples sequenced here, in recent surveillance for nirsevimab resistance-associated mutations^15,40^, and in the wider set of samples from GISAID does not indicate any SNVs restricted to these Boston genomes alone. Thus, we see no evidence of Boston-circulating RSV accumulating treatment-resistant mutations.

There are limitations to our study. First, data comes from a single center. Further surveillance data from other surrounding and not contiguous medical centers is needed to confirm the generalizability of our observations. Second, we were unable to sequence all RSV detected within the BMC community. The samples not sequenced could have a different proportion of RSV-A and RSV-B especially because of the demographic differences. Finally, we can speculate but cannot determine the reasons for the predominance of RSV-B over RSV-A in 2024 as compared to 2022.

This study highlights the importance of regular genomic surveillance to monitor pathogen dynamics. Just two years after the most recent RSV surveillance in Boston^16^, we identified a drastic shift in subtype prevalence in RSV circulating in this area. We also identified differences in transmission dynamics between the subtypes. We saw a variety of RSV-A strains introduced but not expanded over time, whereas we observed clustered transmission of RSV-B and suggestions of sustained RSV-B transmission over the same period. It is tempting to speculate that a previous period of strong RSV-A transmission could create an immunological backdrop that would favor RSV-B transmission. Genomic surveillance of RSV will remain important as new interventions become more common, to monitor for selective pressure and changes in transmission dynamics.

## Acknowledgements

We gratefully acknowledge all data contributors, i.e., the Authors and their Originating laboratories responsible for obtaining the specimens, and their Submitting laboratories for generating the genetic sequence and metadata and sharing via the GISAID Initiative, on which the global comparison and phylogeny of this research is based. Sample and data collection was funded by the Massachusetts Consortium for Pathogen Readiness (Mass-CPR). This publication was supported by the Office of Advanced Molecular Detection, Centers for Disease Control and Prevention through Cooperative Agreement Number CK22-2204. Its contents are solely the responsibility of the authors and do not necessarily represent the official views of the Centers for Disease Control and Prevention. We thank Joseline Velasquez-Reyes and Michelle Nguyen for helpful comments and discussion.

## Data Sharing Statement

Raw sequencing data is available from the Sequencing Read Archive (SRA), within BioProject PRJNA1223358. Genome consensus sequences are available from GISAID (accession numbers EPI_ISL_19725019 to EPI_ISL_19725072). Code used for analysis can be found at github.com/neidl-connor-lab/rsv-2024.

