## Supplemental Information for "Genomic Epidemiology of Respiratory Syncytial Virus in a New England Hospital System, 2024"

Affiliations:
1. National Emerging Infectious Diseases Laboratories, Boston University
2. Department of Virology, Immunology and Microbiology, Boston University Chobanian & Avedisian School of Medicine
3. Department of Medicine, Boston University Chobanian & Avedisian School of Medicine

* These authors contributed equally to this work.

### Table of Contents:

- Supplemental Figure 1 - page 2
- Supplemental Figure 2 - pages 3-4
- Supplemental Figure 3 - pages 5-6
- Supplemental Table 1 - pages 7-8
- Supplemental Table 2 - pages 9-43
- References - page 44

**Supplemental Figure 1**.
Distributions of patient age and sample collection dates stratified by sample subtype.

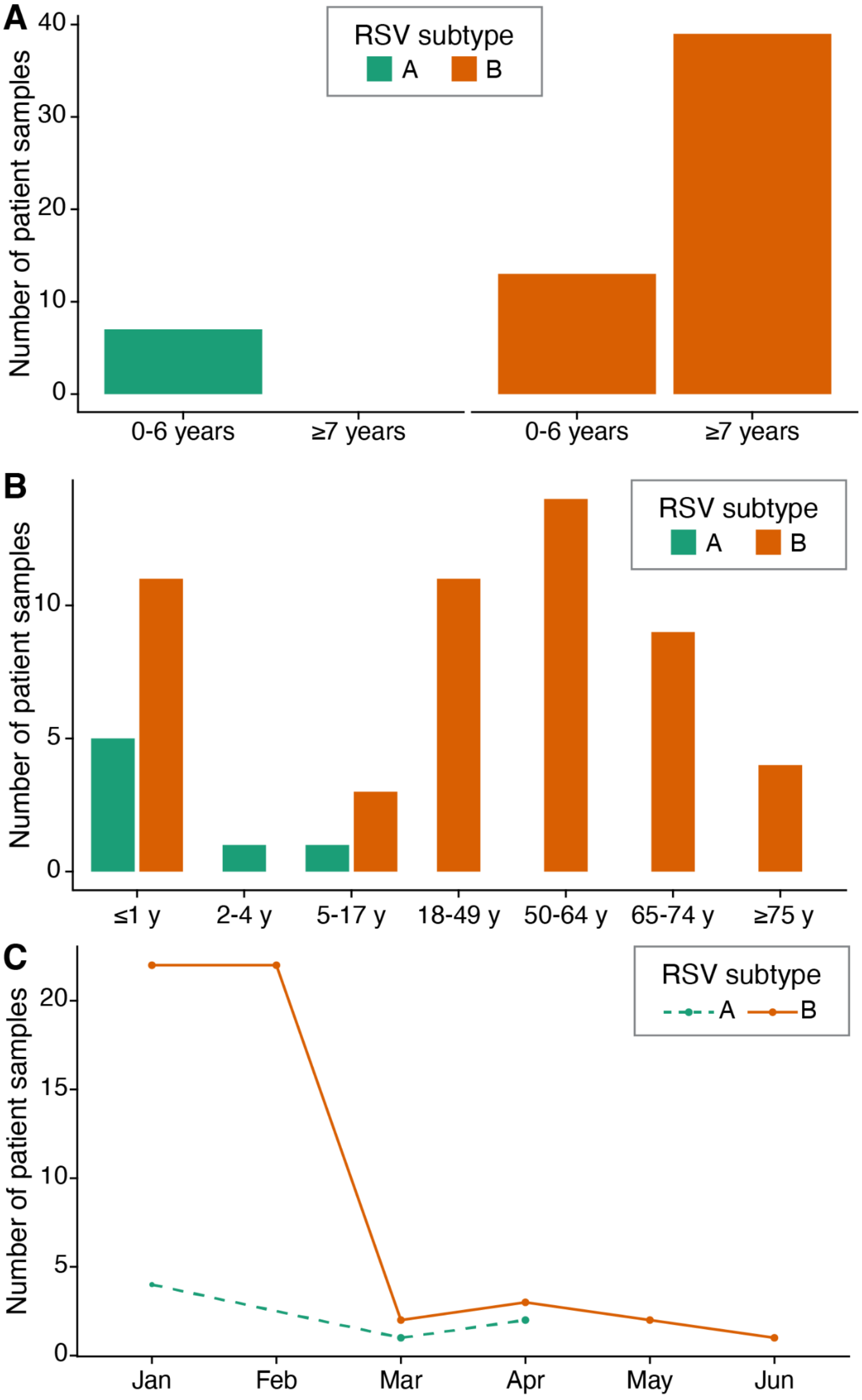


A) Number of samples of each RSV subtype, separated by patient age group. RSV-A samples are in teal, and RSV-B samples are in orange. B) Number of samples of each RSV subtype, separated into a larger number of age groups than in panel A. RSV-A samples are again in teal, and RSV-B samples are in orange. C) Number of samples of each RSV subtype, separated by month of sample collection. RSV-A samples are plotted in teal, with a dashed line, and RSV-B samples are plotted in orange, with a solid line.

**Supplemental Figure 2.**
Comparison of RSV-A genomes from this study and globally.

**A**

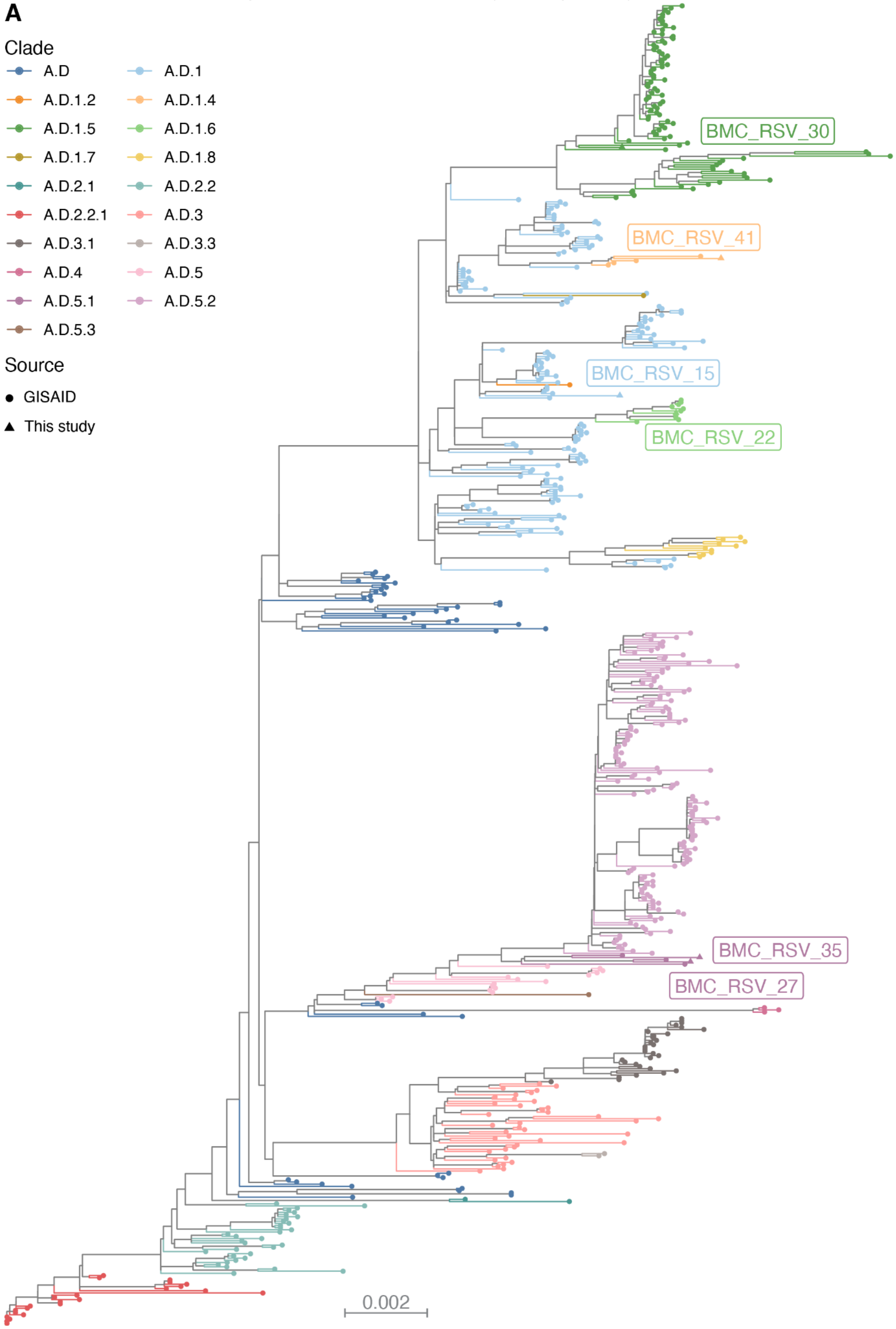

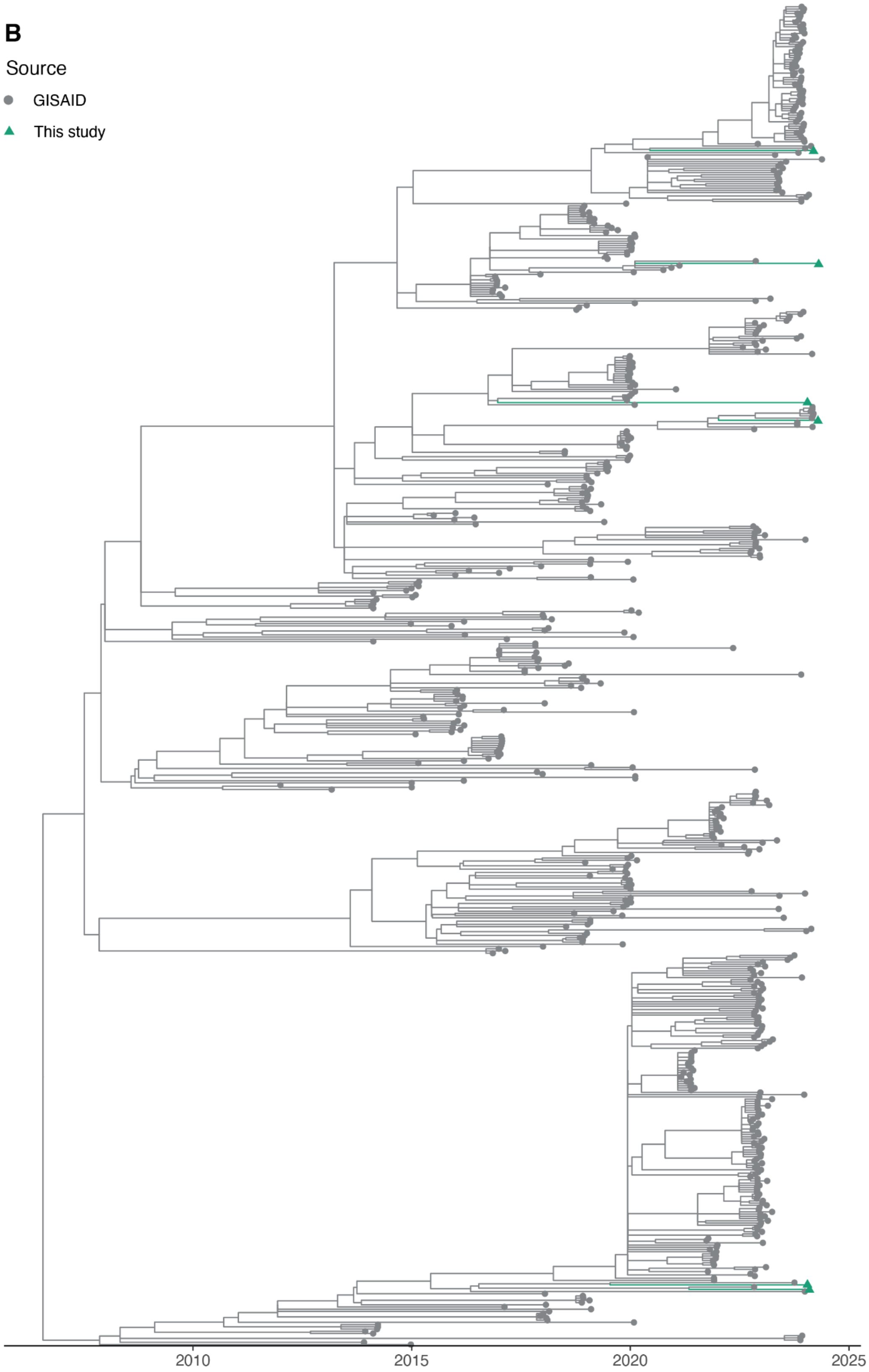


A) Maximum likelihood phylogenetic tree of all RSV-A genomes sequenced in this study (n=6, each labeled with sample ID) and all high coverage, complete RSV-A genomes available in GISAID as of November 8, 2024 (n=600). All samples are colored by clade, as assigned by Nextclade. B) Time-resolved tree of the same genomes in panel A. RSV-A genomes from this study are colored in teal, with triangular tips, and genomes from GISAID are colored in gray, with circular tips.

**Supplemental Figure 3.**
Time-resolved tree of RSV-B samples.

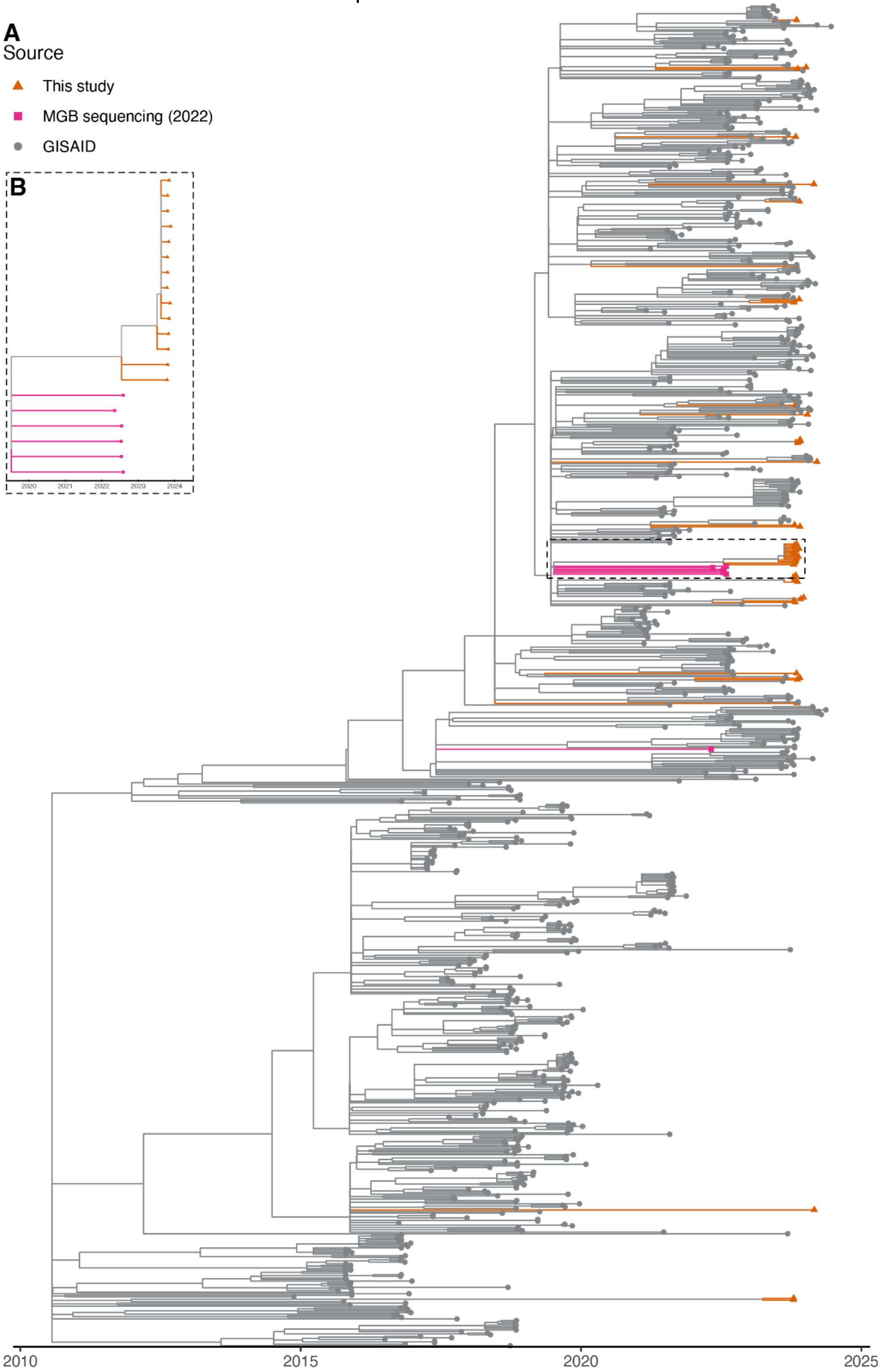


A) Time-resolved tree of RSV-B genomes. Genomes from this study are colored in orange with triangular tips. Genomes from the 2022 sequencing in Boston[1] are colored in pink, with square tips. Genomes from GISAID are colored in gray, with circular tips. The dashed box within the tree outlines the area that is expanded in panel B. B) Expansion of the tree region within the dashed box in panel A. Colors and shapes are the same.

**Supplemental Table 1**. Non-synonymous mutations within RSV-B F gene in Boston and global 2024 genomes.

| Substitution | GISAID 2024 samples | | BMC 2024 samples | | |
|---|---|---|---|---|---|
| | Number | Percent | Number | Percent | Within-sample % (BMC_RSV_14) |
| S8T | 2 | 1.8 | | | |
| F12I | 15 | 13.5 | | | |
| A16T | 1 | 0.9 | | | |
| A19T | 15 | 13.5 | | | |
| L22F | 1 | 0.9 | | | |
| R42K | 44 | 39.6 | 2 | 4.2 | |
| T67N | 1 | 0.9 | | | |
| V103I | 3 | 2.7 | | | |
| E110G | | | 1 | 2.1 | |
| A111T | 1 | 0.9 | | | |
| Y114H | | | 1 | 2.1 | |
| T118A | 9 | 8.1 | | | |
| T122I | 2 | 1.8 | | | |
| V144I | 2 | 1.8 | | | |
| S190N | 100 | 90.1 | 45 | 93.8 | 100.0% |
| R191K | | | 2 | 4.2 | |
| R209Q | | | 3 | 6.2 | |
| S211N | 101 | 91 | 46 | 95.8 | 100.0% |
| S276N | 1 | 0.9 | | | |
| E294G | 2 | 1.8 | | | |
| T324I | 1 | 0.9 | | | |
| K327N | | | 1 | 2.1 | |
| A355T | 1 | 0.9 | | | |
| V360A | 1 | 0.9 | | | |
| D385N | 1 | 0.9 | | | |
| I386M | | | 1 | 2.1 | |
| S389P | 110 | 99.1 | 44 | 91.7 | 99.9% |
| I432V | | | 1 | 2.1 | |

| | | | | | |
|---|---|---|---|---|---|
| K445R | | | 1 | 2.1 | |
| L467F | 1 | 0.9 | 1 | 2.1 | 22.1% |
| R508K | 1 | 0.9 | | | |
| H514R | 1 | 0.9 | | | |
| A529V | | | 6 | 12.5 | 33.0% |
| T558A | | | 1 | 2.1 | 22.1% |

All non-synonymous single nucleotide variants (SNVs) within the F gene of RSV-B. Each row contains one SNV, the number and percentage of samples from GISAID in 2024 that contain it and the number and percentage of genomes in this study that contain it. The final column denotes the within-sample frequency of the variant within sample BMC_RSV_14 in this study, which originated from an individual who received nirsevimab 40 days prior to sample collection. All SNVs above 10% frequency within that sample are included here.

**Supplemental Table 2.**
GISAID and Genbank IDs for genomes used in phylogenetic analyses.

| ID / Accession | Source | RSV Subtype | Country | Year |
|---|---|---|---|---|
| EPI_ISL_19500944 | GISAID | RSV-A | Argentina | 2024 |
| EPI_ISL_19500946 | GISAID | RSV-A | Argentina | 2023 |
| EPI_ISL_19500948 | GISAID | RSV-A | Argentina | 2023 |
| EPI_ISL_19500950 | GISAID | RSV-A | Argentina | 2023 |
| EPI_ISL_19500951 | GISAID | RSV-A | Argentina | 2023 |
| EPI_ISL_19500955 | GISAID | RSV-A | Argentina | 2023 |
| EPI_ISL_19500956 | GISAID | RSV-A | Argentina | 2023 |
| EPI_ISL_19500958 | GISAID | RSV-A | Argentina | 2023 |
| EPI_ISL_19500959 | GISAID | RSV-A | Argentina | 2023 |
| EPI_ISL_19500960 | GISAID | RSV-A | Argentina | 2023 |
| EPI_ISL_19500965 | GISAID | RSV-A | Argentina | 2023 |
| EPI_ISL_19500968 | GISAID | RSV-A | Argentina | 2023 |
| EPI_ISL_19500970 | GISAID | RSV-A | Argentina | 2023 |
| EPI_ISL_19500972 | GISAID | RSV-A | Argentina | 2023 |
| EPI_ISL_19500973 | GISAID | RSV-A | Argentina | 2023 |
| EPI_ISL_19500975 | GISAID | RSV-A | Argentina | 2023 |
| EPI_ISL_19500976 | GISAID | RSV-A | Argentina | 2023 |
| EPI_ISL_19500977 | GISAID | RSV-A | Argentina | 2023 |
| EPI_ISL_19005678 | GISAID | RSV-A | Australia | 2023 |
| EPI_ISL_19005625 | GISAID | RSV-A | Australia | 2023 |
| EPI_ISL_19005634 | GISAID | RSV-A | Australia | 2022 |
| EPI_ISL_19015704 | GISAID | RSV-A | Australia | 2023 |
| EPI_ISL_19048220 | GISAID | RSV-A | Australia | 2019 |
| EPI_ISL_19048575 | GISAID | RSV-A | Australia | 2018 |
| EPI_ISL_19048851 | GISAID | RSV-A | Australia | 2018 |
| EPI_ISL_19049203 | GISAID | RSV-A | Australia | 2018 |
| EPI_ISL_2991498 | GISAID | RSV-A | Australia | 2021 |
| EPI_ISL_18979752 | GISAID | RSV-A | Australia | 2022 |
| EPI_ISL_19048682 | GISAID | RSV-A | Australia | 2018 |
| EPI_ISL_19048021 | GISAID | RSV-A | Brazil | 2019 |

| | | | | |
|---|---|---|---|---|
| EPI_ISL_19048904 | GISAID | RSV-A | Brazil | 2019 |
| EPI_ISL_18999150 | GISAID | RSV-A | Brazil | 2020 |
| EPI_ISL_18999152 | GISAID | RSV-A | Brazil | 2020 |
| EPI_ISL_18999157 | GISAID | RSV-A | Brazil | 2021 |
| EPI_ISL_19047998 | GISAID | RSV-A | Brazil | 2018 |
| EPI_ISL_19048214 | GISAID | RSV-A | Brazil | 2019 |
| EPI_ISL_19048503 | GISAID | RSV-A | Brazil | 2019 |
| EPI_ISL_19048507 | GISAID | RSV-A | Brazil | 2018 |
| EPI_ISL_19048585 | GISAID | RSV-A | Brazil | 2018 |
| EPI_ISL_19048741 | GISAID | RSV-A | Brazil | 2019 |
| EPI_ISL_19048804 | GISAID | RSV-A | Brazil | 2019 |
| EPI_ISL_19048862 | GISAID | RSV-A | Brazil | 2019 |
| EPI_ISL_19048920 | GISAID | RSV-A | Brazil | 2019 |
| EPI_ISL_19048203 | GISAID | RSV-A | Brazil | 2019 |
| EPI_ISL_19048415 | GISAID | RSV-A | Brazil | 2019 |
| EPI_ISL_19048798 | GISAID | RSV-A | Brazil | 2019 |
| EPI_ISL_19048886 | GISAID | RSV-A | Brazil | 2019 |
| EPI_ISL_19048040 | GISAID | RSV-A | Canada | 2019 |
| EPI_ISL_19048080 | GISAID | RSV-A | Canada | 2019 |
| EPI_ISL_19048093 | GISAID | RSV-A | Canada | 2019 |
| EPI_ISL_19048142 | GISAID | RSV-A | Canada | 2020 |
| EPI_ISL_19048180 | GISAID | RSV-A | Canada | 2019 |
| EPI_ISL_19048251 | GISAID | RSV-A | Canada | 2020 |
| EPI_ISL_19048375 | GISAID | RSV-A | Canada | 2019 |
| EPI_ISL_19048386 | GISAID | RSV-A | Canada | 2019 |
| EPI_ISL_19048388 | GISAID | RSV-A | Canada | 2019 |
| EPI_ISL_19048496 | GISAID | RSV-A | Canada | 2019 |
| EPI_ISL_19048916 | GISAID | RSV-A | Canada | 2019 |
| EPI_ISL_19049078 | GISAID | RSV-A | Canada | 2018 |
| EPI_ISL_19049097 | GISAID | RSV-A | Canada | 2019 |
| EPI_ISL_19049162 | GISAID | RSV-A | Canada | 2018 |
| EPI_ISL_19049185 | GISAID | RSV-A | Canada | 2019 |

| | | | | |
|---|---|---|---|---|
| EPI_ISL_19049186 | GISAID | RSV-A | Canada | 2018 |
| EPI_ISL_19049194 | GISAID | RSV-A | Canada | 2020 |
| EPI_ISL_19049196 | GISAID | RSV-A | Canada | 2019 |
| EPI_ISL_19049265 | GISAID | RSV-A | Canada | 2019 |
| EPI_ISL_19048117 | GISAID | RSV-A | Canada | 2019 |
| EPI_ISL_19048335 | GISAID | RSV-A | Canada | 2019 |
| EPI_ISL_19048359 | GISAID | RSV-A | Canada | 2019 |
| EPI_ISL_19048610 | GISAID | RSV-A | Canada | 2020 |
| EPI_ISL_19048942 | GISAID | RSV-A | Canada | 2019 |
| EPI_ISL_19048315 | GISAID | RSV-A | Canada | 2019 |
| EPI_ISL_19048402 | GISAID | RSV-A | Canada | 2018 |
| EPI_ISL_19048551 | GISAID | RSV-A | Canada | 2019 |
| EPI_ISL_19048561 | GISAID | RSV-A | Canada | 2018 |
| EPI_ISL_19048786 | GISAID | RSV-A | Canada | 2019 |
| EPI_ISL_18810665 | GISAID | RSV-A | Canary Islands | 2022 |
| EPI_ISL_18810669 | GISAID | RSV-A | Canary Islands | 2022 |
| EPI_ISL_18698561 | GISAID | RSV-A | Canary Islands | 2023 |
| EPI_ISL_18698562 | GISAID | RSV-A | Canary Islands | 2023 |
| EPI_ISL_18698563 | GISAID | RSV-A | Canary Islands | 2023 |
| EPI_ISL_18698564 | GISAID | RSV-A | Canary Islands | 2023 |
| EPI_ISL_18698565 | GISAID | RSV-A | Canary Islands | 2023 |
| EPI_ISL_18698567 | GISAID | RSV-A | Canary Islands | 2023 |
| EPI_ISL_18698569 | GISAID | RSV-A | Canary Islands | 2023 |
| EPI_ISL_18698571 | GISAID | RSV-A | Canary Islands | 2023 |
| EPI_ISL_18698572 | GISAID | RSV-A | Canary Islands | 2023 |
| EPI_ISL_18698575 | GISAID | RSV-A | Canary Islands | 2023 |
| EPI_ISL_18698576 | GISAID | RSV-A | Canary Islands | 2023 |
| EPI_ISL_18698579 | GISAID | RSV-A | Canary Islands | 2023 |
| EPI_ISL_18698582 | GISAID | RSV-A | Canary Islands | 2023 |
| EPI_ISL_18698583 | GISAID | RSV-A | Canary Islands | 2023 |
| EPI_ISL_18698584 | GISAID | RSV-A | Canary Islands | 2023 |
| EPI_ISL_18698586 | GISAID | RSV-A | Canary Islands | 2023 |

| | | | | |
|---|---|---|---|---|
| EPI_ISL_18698587 | GISAID | RSV-A | Canary Islands | 2023 |
| EPI_ISL_18698589 | GISAID | RSV-A | Canary Islands | 2023 |
| EPI_ISL_18698602 | GISAID | RSV-A | Canary Islands | 2022 |
| EPI_ISL_18698616 | GISAID | RSV-A | Canary Islands | 2022 |
| EPI_ISL_18698682 | GISAID | RSV-A | Canary Islands | 2023 |
| EPI_ISL_18698683 | GISAID | RSV-A | Canary Islands | 2023 |
| EPI_ISL_18698684 | GISAID | RSV-A | Canary Islands | 2023 |
| EPI_ISL_18698687 | GISAID | RSV-A | Canary Islands | 2023 |
| EPI_ISL_18698690 | GISAID | RSV-A | Canary Islands | 2023 |
| EPI_ISL_18698695 | GISAID | RSV-A | Canary Islands | 2023 |
| EPI_ISL_18698701 | GISAID | RSV-A | Canary Islands | 2023 |
| EPI_ISL_18698703 | GISAID | RSV-A | Canary Islands | 2023 |
| EPI_ISL_18698707 | GISAID | RSV-A | Canary Islands | 2023 |
| EPI_ISL_18698708 | GISAID | RSV-A | Canary Islands | 2023 |
| EPI_ISL_18698709 | GISAID | RSV-A | Canary Islands | 2023 |
| EPI_ISL_18698710 | GISAID | RSV-A | Canary Islands | 2023 |
| EPI_ISL_18698711 | GISAID | RSV-A | Canary Islands | 2023 |
| EPI_ISL_18698713 | GISAID | RSV-A | Canary Islands | 2023 |
| EPI_ISL_18698715 | GISAID | RSV-A | Canary Islands | 2023 |
| EPI_ISL_18698716 | GISAID | RSV-A | Canary Islands | 2023 |
| EPI_ISL_18810656 | GISAID | RSV-A | Canary Islands | 2022 |
| EPI_ISL_18810670 | GISAID | RSV-A | Canary Islands | 2022 |
| EPI_ISL_18810671 | GISAID | RSV-A | Canary Islands | 2022 |
| EPI_ISL_18810672 | GISAID | RSV-A | Canary Islands | 2022 |
| EPI_ISL_18810677 | GISAID | RSV-A | Canary Islands | 2022 |
| EPI_ISL_18810682 | GISAID | RSV-A | Canary Islands | 2023 |
| EPI_ISL_18810683 | GISAID | RSV-A | Canary Islands | 2023 |
| EPI_ISL_18810684 | GISAID | RSV-A | Canary Islands | 2023 |
| EPI_ISL_18810685 | GISAID | RSV-A | Canary Islands | 2023 |
| EPI_ISL_18810686 | GISAID | RSV-A | Canary Islands | 2023 |
| EPI_ISL_18810687 | GISAID | RSV-A | Canary Islands | 2023 |
| EPI_ISL_18810688 | GISAID | RSV-A | Canary Islands | 2023 |

| | | | | |
|---|---|---|---|---|
| EPI_ISL_18810689 | GISAID | RSV-A | Canary Islands | 2023 |
| EPI_ISL_18810690 | GISAID | RSV-A | Canary Islands | 2023 |
| EPI_ISL_18810691 | GISAID | RSV-A | Canary Islands | 2023 |
| EPI_ISL_18810692 | GISAID | RSV-A | Canary Islands | 2023 |
| EPI_ISL_18810693 | GISAID | RSV-A | Canary Islands | 2023 |
| EPI_ISL_18810694 | GISAID | RSV-A | Canary Islands | 2023 |
| EPI_ISL_18810695 | GISAID | RSV-A | Canary Islands | 2023 |
| EPI_ISL_18810696 | GISAID | RSV-A | Canary Islands | 2023 |
| EPI_ISL_18810698 | GISAID | RSV-A | Canary Islands | 2023 |
| EPI_ISL_18810699 | GISAID | RSV-A | Canary Islands | 2023 |
| EPI_ISL_18810700 | GISAID | RSV-A | Canary Islands | 2023 |
| EPI_ISL_18810701 | GISAID | RSV-A | Canary Islands | 2023 |
| EPI_ISL_18810702 | GISAID | RSV-A | Canary Islands | 2023 |
| EPI_ISL_18810703 | GISAID | RSV-A | Canary Islands | 2023 |
| EPI_ISL_18810704 | GISAID | RSV-A | Canary Islands | 2023 |
| EPI_ISL_18810706 | GISAID | RSV-A | Canary Islands | 2023 |
| EPI_ISL_18810707 | GISAID | RSV-A | Canary Islands | 2023 |
| EPI_ISL_18810708 | GISAID | RSV-A | Canary Islands | 2023 |
| EPI_ISL_18810709 | GISAID | RSV-A | Canary Islands | 2023 |
| EPI_ISL_18810710 | GISAID | RSV-A | Canary Islands | 2023 |
| EPI_ISL_18810711 | GISAID | RSV-A | Canary Islands | 2023 |
| EPI_ISL_18698547 | GISAID | RSV-A | Canary Islands | 2023 |
| EPI_ISL_18698557 | GISAID | RSV-A | Canary Islands | 2023 |
| EPI_ISL_18698558 | GISAID | RSV-A | Canary Islands | 2023 |
| EPI_ISL_18698559 | GISAID | RSV-A | Canary Islands | 2023 |
| EPI_ISL_18698649 | GISAID | RSV-A | Canary Islands | 2022 |
| EPI_ISL_18482766 | GISAID | RSV-A | China | 2019 |
| EPI_ISL_18482800 | GISAID | RSV-A | China | 2020 |
| EPI_ISL_18482814 | GISAID | RSV-A | China | 2019 |
| EPI_ISL_18482815 | GISAID | RSV-A | China | 2018 |
| EPI_ISL_19493456 | GISAID | RSV-A | Cote d'Ivoire | 2023 |
| EPI_ISL_19493521 | GISAID | RSV-A | Cote d'Ivoire | 2023 |

| | | | | |
|---|---|---|---|---|
| EPI_ISL_412866 | GISAID | RSV-A | England | 2017 |
| EPI_ISL_1647388 | GISAID | RSV-A | England | 2019 |
| EPI_ISL_18591755 | GISAID | RSV-A | England | 2023 |
| EPI_ISL_412865 | GISAID | RSV-A | England | 2017 |
| EPI_ISL_732338 | GISAID | RSV-A | England | 2017 |
| EPI_ISL_732345 | GISAID | RSV-A | England | 2017 |
| EPI_ISL_732346 | GISAID | RSV-A | England | 2017 |
| EPI_ISL_732361 | GISAID | RSV-A | England | 2018 |
| EPI_ISL_10954007 | GISAID | RSV-A | England | 2017 |
| EPI_ISL_10954008 | GISAID | RSV-A | England | 2017 |
| EPI_ISL_19048004 | GISAID | RSV-A | Finland | 2019 |
| EPI_ISL_19048296 | GISAID | RSV-A | Finland | 2020 |
| EPI_ISL_19049110 | GISAID | RSV-A | Finland | 2019 |
| EPI_ISL_19047982 | GISAID | RSV-A | Finland | 2018 |
| EPI_ISL_19048119 | GISAID | RSV-A | Finland | 2020 |
| EPI_ISL_19016305 | GISAID | RSV-A | France | 2023 |
| EPI_ISL_19016313 | GISAID | RSV-A | France | 2023 |
| EPI_ISL_19048015 | GISAID | RSV-A | France | 2019 |
| EPI_ISL_19048483 | GISAID | RSV-A | France | 2019 |
| EPI_ISL_19049147 | GISAID | RSV-A | France | 2018 |
| EPI_ISL_19049252 | GISAID | RSV-A | France | 2019 |
| EPI_ISL_19048976 | GISAID | RSV-A | France | 2019 |
| EPI_ISL_19048414 | GISAID | RSV-A | France | 2019 |
| EPI_ISL_19048432 | GISAID | RSV-A | France | 2019 |
| EPI_ISL_17995656 | GISAID | RSV-A | Germany | 2017 |
| EPI_ISL_19048245 | GISAID | RSV-A | Germany | 2020 |
| EPI_ISL_19047985 | GISAID | RSV-A | Germany | 2020 |
| EPI_ISL_19048545 | GISAID | RSV-A | Germany | 2019 |
| EPI_ISL_19049032 | GISAID | RSV-A | Italy | 2020 |
| EPI_ISL_19063348 | GISAID | RSV-A | Italy | 2022 |
| EPI_ISL_19063351 | GISAID | RSV-A | Italy | 2023 |
| EPI_ISL_19063357 | GISAID | RSV-A | Italy | 2021 |

| | | | | |
|---|---|---|---|---|
| EPI_ISL_19063358 | GISAID | RSV-A | Italy | 2018 |
| EPI_ISL_19063359 | GISAID | RSV-A | Italy | 2020 |
| EPI_ISL_19063360 | GISAID | RSV-A | Italy | 2018 |
| EPI_ISL_19063363 | GISAID | RSV-A | Italy | 2020 |
| EPI_ISL_19063364 | GISAID | RSV-A | Italy | 2018 |
| EPI_ISL_19063365 | GISAID | RSV-A | Italy | 2021 |
| EPI_ISL_19063373 | GISAID | RSV-A | Italy | 2018 |
| EPI_ISL_19063374 | GISAID | RSV-A | Italy | 2023 |
| EPI_ISL_19063376 | GISAID | RSV-A | Italy | 2021 |
| EPI_ISL_19063390 | GISAID | RSV-A | Italy | 2017 |
| EPI_ISL_19063393 | GISAID | RSV-A | Italy | 2021 |
| EPI_ISL_19063401 | GISAID | RSV-A | Italy | 2018 |
| EPI_ISL_19063403 | GISAID | RSV-A | Italy | 2019 |
| EPI_ISL_19063404 | GISAID | RSV-A | Italy | 2021 |
| EPI_ISL_19063405 | GISAID | RSV-A | Italy | 2021 |
| EPI_ISL_19063408 | GISAID | RSV-A | Italy | 2018 |
| EPI_ISL_19063410 | GISAID | RSV-A | Italy | 2022 |
| EPI_ISL_19063414 | GISAID | RSV-A | Italy | 2020 |
| EPI_ISL_19063415 | GISAID | RSV-A | Italy | 2019 |
| EPI_ISL_19063416 | GISAID | RSV-A | Italy | 2021 |
| EPI_ISL_19063417 | GISAID | RSV-A | Italy | 2021 |
| EPI_ISL_19063420 | GISAID | RSV-A | Italy | 2021 |
| EPI_ISL_18446782 | GISAID | RSV-A | Italy | 2021 |
| EPI_ISL_18446784 | GISAID | RSV-A | Italy | 2021 |
| EPI_ISL_19048237 | GISAID | RSV-A | Japan | 2017 |
| EPI_ISL_413222 | GISAID | RSV-A | Madagascar | 2014 |
| EPI_ISL_413353 | GISAID | RSV-A | Madagascar | 2016 |
| EPI_ISL_15120703 | GISAID | RSV-A | Morocco | 2016 |
| EPI_ISL_15120706 | GISAID | RSV-A | Morocco | 2016 |
| EPI_ISL_15120665 | GISAID | RSV-A | Morocco | 2014 |
| EPI_ISL_15120666 | GISAID | RSV-A | Morocco | 2014 |
| EPI_ISL_15120667 | GISAID | RSV-A | Morocco | 2014 |

| | | | | |
|---|---|---|---|---|
| EPI_ISL_15120668 | GISAID | RSV-A | Morocco | 2014 |
| EPI_ISL_15120669 | GISAID | RSV-A | Morocco | 2014 |
| EPI_ISL_15120713 | GISAID | RSV-A | Morocco | 2016 |
| EPI_ISL_15120715 | GISAID | RSV-A | Morocco | 2016 |
| EPI_ISL_15120716 | GISAID | RSV-A | Morocco | 2016 |
| EPI_ISL_15120717 | GISAID | RSV-A | Morocco | 2016 |
| EPI_ISL_15120719 | GISAID | RSV-A | Morocco | 2016 |
| EPI_ISL_15120721 | GISAID | RSV-A | Morocco | 2016 |
| EPI_ISL_15120723 | GISAID | RSV-A | Morocco | 2016 |
| EPI_ISL_15120726 | GISAID | RSV-A | Morocco | 2016 |
| EPI_ISL_15120727 | GISAID | RSV-A | Morocco | 2016 |
| EPI_ISL_15120729 | GISAID | RSV-A | Morocco | 2016 |
| EPI_ISL_15120730 | GISAID | RSV-A | Morocco | 2016 |
| EPI_ISL_15120738 | GISAID | RSV-A | Morocco | 2017 |
| EPI_ISL_15120739 | GISAID | RSV-A | Morocco | 2017 |
| EPI_ISL_15120743 | GISAID | RSV-A | Morocco | 2017 |
| EPI_ISL_15120745 | GISAID | RSV-A | Morocco | 2017 |
| EPI_ISL_15120770 | GISAID | RSV-A | Morocco | 2017 |
| EPI_ISL_15120771 | GISAID | RSV-A | Morocco | 2017 |
| EPI_ISL_15120772 | GISAID | RSV-A | Morocco | 2017 |
| EPI_ISL_15120773 | GISAID | RSV-A | Morocco | 2017 |
| EPI_ISL_15120774 | GISAID | RSV-A | Morocco | 2017 |
| EPI_ISL_15120775 | GISAID | RSV-A | Morocco | 2017 |
| EPI_ISL_15120776 | GISAID | RSV-A | Morocco | 2017 |
| EPI_ISL_15120778 | GISAID | RSV-A | Morocco | 2017 |
| EPI_ISL_15120780 | GISAID | RSV-A | Morocco | 2018 |
| EPI_ISL_15120781 | GISAID | RSV-A | Morocco | 2018 |
| EPI_ISL_15120782 | GISAID | RSV-A | Morocco | 2018 |
| EPI_ISL_15120783 | GISAID | RSV-A | Morocco | 2018 |
| EPI_ISL_15120785 | GISAID | RSV-A | Morocco | 2019 |
| EPI_ISL_15120786 | GISAID | RSV-A | Morocco | 2019 |
| EPI_ISL_15120670 | GISAID | RSV-A | Morocco | 2014 |

| | | | | |
|---|---|---|---|---|
| EPI_ISL_15120671 | GISAID | RSV-A | Morocco | 2014 |
| EPI_ISL_15120672 | GISAID | RSV-A | Morocco | 2014 |
| EPI_ISL_15120674 | GISAID | RSV-A | Morocco | 2014 |
| EPI_ISL_15120675 | GISAID | RSV-A | Morocco | 2014 |
| EPI_ISL_15120676 | GISAID | RSV-A | Morocco | 2014 |
| EPI_ISL_15120677 | GISAID | RSV-A | Morocco | 2014 |
| EPI_ISL_15120678 | GISAID | RSV-A | Morocco | 2015 |
| EPI_ISL_15120679 | GISAID | RSV-A | Morocco | 2015 |
| EPI_ISL_15120680 | GISAID | RSV-A | Morocco | 2015 |
| EPI_ISL_15120681 | GISAID | RSV-A | Morocco | 2015 |
| EPI_ISL_15120682 | GISAID | RSV-A | Morocco | 2015 |
| EPI_ISL_15120683 | GISAID | RSV-A | Morocco | 2015 |
| EPI_ISL_15120688 | GISAID | RSV-A | Morocco | 2015 |
| EPI_ISL_15120689 | GISAID | RSV-A | Morocco | 2015 |
| EPI_ISL_15120692 | GISAID | RSV-A | Morocco | 2016 |
| EPI_ISL_15120694 | GISAID | RSV-A | Morocco | 2016 |
| EPI_ISL_19048226 | GISAID | RSV-A | Netherlands | 2017 |
| EPI_ISL_19048510 | GISAID | RSV-A | Netherlands | 2017 |
| EPI_ISL_19206528 | GISAID | RSV-A | New_Zealand | 2017 |
| EPI_ISL_19207121 | GISAID | RSV-A | New_Zealand | 2017 |
| EPI_ISL_412458 | GISAID | RSV-A | Nicaragua | 2016 |
| EPI_ISL_19048062 | GISAID | RSV-A | Russia | 2020 |
| EPI_ISL_19047978 | GISAID | RSV-A | Russia | 2020 |
| EPI_ISL_19048641 | GISAID | RSV-A | Russia | 2020 |
| EPI_ISL_19048775 | GISAID | RSV-A | Russia | 2019 |
| EPI_ISL_19442427 | GISAID | RSV-A | Scotland | 2022 |
| EPI_ISL_19442449 | GISAID | RSV-A | Scotland | 2019 |
| EPI_ISL_19442572 | GISAID | RSV-A | Scotland | 2022 |
| EPI_ISL_19442613 | GISAID | RSV-A | Scotland | 2023 |
| EPI_ISL_19442637 | GISAID | RSV-A | Scotland | 2022 |
| EPI_ISL_19442701 | GISAID | RSV-A | Scotland | 2022 |
| EPI_ISL_19442707 | GISAID | RSV-A | Scotland | 2019 |

| | | | | |
|---|---|---|---|---|
| EPI_ISL_19442399 | GISAID | RSV-A | Scotland | 2022 |
| EPI_ISL_19442488 | GISAID | RSV-A | Scotland | 2020 |
| EPI_ISL_19049279 | GISAID | RSV-A | South Africa | 2018 |
| EPI_ISL_19220723 | GISAID | RSV-A | South Africa | 2023 |
| EPI_ISL_19220725 | GISAID | RSV-A | South Africa | 2023 |
| EPI_ISL_19220726 | GISAID | RSV-A | South Africa | 2023 |
| EPI_ISL_19090981 | GISAID | RSV-A | South Korea | 2021 |
| EPI_ISL_19090982 | GISAID | RSV-A | South Korea | 2021 |
| EPI_ISL_19090983 | GISAID | RSV-A | South Korea | 2021 |
| EPI_ISL_19090984 | GISAID | RSV-A | South Korea | 2021 |
| EPI_ISL_19090985 | GISAID | RSV-A | South Korea | 2021 |
| EPI_ISL_19090986 | GISAID | RSV-A | South Korea | 2022 |
| EPI_ISL_19090987 | GISAID | RSV-A | South Korea | 2022 |
| EPI_ISL_19090988 | GISAID | RSV-A | South Korea | 2022 |
| EPI_ISL_19090989 | GISAID | RSV-A | South Korea | 2022 |
| EPI_ISL_19090990 | GISAID | RSV-A | South Korea | 2022 |
| EPI_ISL_19090991 | GISAID | RSV-A | South Korea | 2022 |
| EPI_ISL_19090992 | GISAID | RSV-A | South Korea | 2022 |
| EPI_ISL_19090993 | GISAID | RSV-A | South Korea | 2022 |
| EPI_ISL_19090996 | GISAID | RSV-A | South Korea | 2022 |
| EPI_ISL_19090997 | GISAID | RSV-A | South Korea | 2022 |
| EPI_ISL_19090998 | GISAID | RSV-A | South Korea | 2022 |
| EPI_ISL_19090999 | GISAID | RSV-A | South Korea | 2022 |
| EPI_ISL_19048548 | GISAID | RSV-A | South Korea | 2020 |
| EPI_ISL_19091000 | GISAID | RSV-A | South Korea | 2023 |
| EPI_ISL_19091003 | GISAID | RSV-A | South Korea | 2023 |
| EPI_ISL_19091004 | GISAID | RSV-A | South Korea | 2023 |
| EPI_ISL_18682141 | GISAID | RSV-A | Spain | 2021 |
| EPI_ISL_18682142 | GISAID | RSV-A | Spain | 2021 |
| EPI_ISL_18682143 | GISAID | RSV-A | Spain | 2021 |
| EPI_ISL_18682147 | GISAID | RSV-A | Spain | 2021 |
| EPI_ISL_18682212 | GISAID | RSV-A | Spain | 2022 |

| | | | | |
|---|---|---|---|---|
| EPI_ISL_18682214 | GISAID | RSV-A | Spain | 2022 |
| EPI_ISL_18682216 | GISAID | RSV-A | Spain | 2022 |
| EPI_ISL_18682267 | GISAID | RSV-A | Spain | 2015 |
| EPI_ISL_18682268 | GISAID | RSV-A | Spain | 2015 |
| EPI_ISL_18682269 | GISAID | RSV-A | Spain | 2016 |
| EPI_ISL_18682309 | GISAID | RSV-A | Spain | 2020 |
| EPI_ISL_18682315 | GISAID | RSV-A | Spain | 2013 |
| EPI_ISL_18682316 | GISAID | RSV-A | Spain | 2014 |
| EPI_ISL_18682317 | GISAID | RSV-A | Spain | 2015 |
| EPI_ISL_18682323 | GISAID | RSV-A | Spain | 2017 |
| EPI_ISL_18682332 | GISAID | RSV-A | Spain | 2018 |
| EPI_ISL_18682336 | GISAID | RSV-A | Spain | 2019 |
| EPI_ISL_18682337 | GISAID | RSV-A | Spain | 2019 |
| EPI_ISL_18682389 | GISAID | RSV-A | Spain | 2021 |
| EPI_ISL_18682401 | GISAID | RSV-A | Spain | 2021 |
| EPI_ISL_18682412 | GISAID | RSV-A | Spain | 2021 |
| EPI_ISL_18682420 | GISAID | RSV-A | Spain | 2017 |
| EPI_ISL_18682421 | GISAID | RSV-A | Spain | 2019 |
| EPI_ISL_18682425 | GISAID | RSV-A | Spain | 2020 |
| EPI_ISL_18682434 | GISAID | RSV-A | Spain | 2021 |
| EPI_ISL_18682439 | GISAID | RSV-A | Spain | 2016 |
| EPI_ISL_18682443 | GISAID | RSV-A | Spain | 2014 |
| EPI_ISL_18682446 | GISAID | RSV-A | Spain | 2021 |
| EPI_ISL_18746149 | GISAID | RSV-A | Spain | 2023 |
| EPI_ISL_18928402 | GISAID | RSV-A | Spain | 2023 |
| EPI_ISL_19048049 | GISAID | RSV-A | Spain | 2019 |
| EPI_ISL_19048908 | GISAID | RSV-A | Spain | 2019 |
| EPI_ISL_19049009 | GISAID | RSV-A | Spain | 2019 |
| EPI_ISL_19049232 | GISAID | RSV-A | Spain | 2018 |
| EPI_ISL_19131452 | GISAID | RSV-A | Spain | 2022 |
| EPI_ISL_19131453 | GISAID | RSV-A | Spain | 2022 |
| EPI_ISL_19131454 | GISAID | RSV-A | Spain | 2022 |

| | | | | |
|---|---|---|---|---|
| EPI_ISL_19131455 | GISAID | RSV-A | Spain | 2022 |
| EPI_ISL_19131456 | GISAID | RSV-A | Spain | 2022 |
| EPI_ISL_19131459 | GISAID | RSV-A | Spain | 2022 |
| EPI_ISL_19131467 | GISAID | RSV-A | Spain | 2022 |
| EPI_ISL_19131478 | GISAID | RSV-A | Spain | 2022 |
| EPI_ISL_19131486 | GISAID | RSV-A | Spain | 2022 |
| EPI_ISL_19131492 | GISAID | RSV-A | Spain | 2022 |
| EPI_ISL_19131494 | GISAID | RSV-A | Spain | 2022 |
| EPI_ISL_19131496 | GISAID | RSV-A | Spain | 2022 |
| EPI_ISL_19131497 | GISAID | RSV-A | Spain | 2022 |
| EPI_ISL_19131498 | GISAID | RSV-A | Spain | 2022 |
| EPI_ISL_19131564 | GISAID | RSV-A | Spain | 2022 |
| EPI_ISL_19131565 | GISAID | RSV-A | Spain | 2022 |
| EPI_ISL_19131571 | GISAID | RSV-A | Spain | 2022 |
| EPI_ISL_19131574 | GISAID | RSV-A | Spain | 2022 |
| EPI_ISL_19131578 | GISAID | RSV-A | Spain | 2022 |
| EPI_ISL_19131579 | GISAID | RSV-A | Spain | 2022 |
| EPI_ISL_19131582 | GISAID | RSV-A | Spain | 2022 |
| EPI_ISL_19131589 | GISAID | RSV-A | Spain | 2023 |
| EPI_ISL_19131591 | GISAID | RSV-A | Spain | 2023 |
| EPI_ISL_19131592 | GISAID | RSV-A | Spain | 2023 |
| EPI_ISL_19131594 | GISAID | RSV-A | Spain | 2023 |
| EPI_ISL_19131597 | GISAID | RSV-A | Spain | 2023 |
| EPI_ISL_18682101 | GISAID | RSV-A | Spain | 2021 |
| EPI_ISL_18682103 | GISAID | RSV-A | Spain | 2021 |
| EPI_ISL_18682105 | GISAID | RSV-A | Spain | 2021 |
| EPI_ISL_18682107 | GISAID | RSV-A | Spain | 2021 |
| EPI_ISL_18682111 | GISAID | RSV-A | Spain | 2021 |
| EPI_ISL_18682114 | GISAID | RSV-A | Spain | 2021 |
| EPI_ISL_18682118 | GISAID | RSV-A | Spain | 2021 |
| EPI_ISL_18682127 | GISAID | RSV-A | Spain | 2023 |
| EPI_ISL_18682128 | GISAID | RSV-A | Spain | 2013 |

| | | | | |
|---|---|---|---|---|
| EPI_ISL_18682137 | GISAID | RSV-A | Spain | 2021 |
| EPI_ISL_18682145 | GISAID | RSV-A | Spain | 2021 |
| EPI_ISL_18682150 | GISAID | RSV-A | Spain | 2021 |
| EPI_ISL_18682152 | GISAID | RSV-A | Spain | 2021 |
| EPI_ISL_18682153 | GISAID | RSV-A | Spain | 2023 |
| EPI_ISL_18682161 | GISAID | RSV-A | Spain | 2021 |
| EPI_ISL_18682162 | GISAID | RSV-A | Spain | 2021 |
| EPI_ISL_18682169 | GISAID | RSV-A | Spain | 2023 |
| EPI_ISL_18682170 | GISAID | RSV-A | Spain | 2018 |
| EPI_ISL_18682171 | GISAID | RSV-A | Spain | 2021 |
| EPI_ISL_18682175 | GISAID | RSV-A | Spain | 2021 |
| EPI_ISL_18682178 | GISAID | RSV-A | Spain | 2022 |
| EPI_ISL_18682181 | GISAID | RSV-A | Spain | 2022 |
| EPI_ISL_18682225 | GISAID | RSV-A | Spain | 2015 |
| EPI_ISL_18682234 | GISAID | RSV-A | Spain | 2016 |
| EPI_ISL_18682238 | GISAID | RSV-A | Spain | 2017 |
| EPI_ISL_18682241 | GISAID | RSV-A | Spain | 2019 |
| EPI_ISL_18682242 | GISAID | RSV-A | Spain | 2019 |
| EPI_ISL_18682253 | GISAID | RSV-A | Spain | 2021 |
| EPI_ISL_18682271 | GISAID | RSV-A | Spain | 2016 |
| EPI_ISL_18682278 | GISAID | RSV-A | Spain | 2016 |
| EPI_ISL_18682282 | GISAID | RSV-A | Spain | 2017 |
| EPI_ISL_18682293 | GISAID | RSV-A | Spain | 2018 |
| EPI_ISL_18682294 | GISAID | RSV-A | Spain | 2018 |
| EPI_ISL_18682296 | GISAID | RSV-A | Spain | 2019 |
| EPI_ISL_18682297 | GISAID | RSV-A | Spain | 2019 |
| EPI_ISL_18682347 | GISAID | RSV-A | Spain | 2016 |
| EPI_ISL_18682348 | GISAID | RSV-A | Spain | 2016 |
| EPI_ISL_18682349 | GISAID | RSV-A | Spain | 2016 |
| EPI_ISL_18682350 | GISAID | RSV-A | Spain | 2016 |
| EPI_ISL_18682354 | GISAID | RSV-A | Spain | 2018 |
| EPI_ISL_18682364 | GISAID | RSV-A | Spain | 2022 |

| | | | | |
|---|---|---|---|---|
| EPI_ISL_18682366 | GISAID | RSV-A | Spain | 2015 |
| EPI_ISL_18682367 | GISAID | RSV-A | Spain | 2016 |
| EPI_ISL_18682370 | GISAID | RSV-A | Spain | 2017 |
| EPI_ISL_18682386 | GISAID | RSV-A | Spain | 2018 |
| EPI_ISL_18968128 | GISAID | RSV-A | Spain | 2023 |
| EPI_ISL_19048617 | GISAID | RSV-A | Spain | 2019 |
| EPI_ISL_19048815 | GISAID | RSV-A | Spain | 2018 |
| EPI_ISL_19048872 | GISAID | RSV-A | Spain | 2020 |
| EPI_ISL_19048968 | GISAID | RSV-A | Spain | 2019 |
| EPI_ISL_19048995 | GISAID | RSV-A | Spain | 2018 |
| EPI_ISL_19049205 | GISAID | RSV-A | Spain | 2020 |
| EPI_ISL_19131520 | GISAID | RSV-A | Spain | 2022 |
| EPI_ISL_19131524 | GISAID | RSV-A | Spain | 2022 |
| EPI_ISL_19131528 | GISAID | RSV-A | Spain | 2022 |
| EPI_ISL_19131529 | GISAID | RSV-A | Spain | 2022 |
| EPI_ISL_19131535 | GISAID | RSV-A | Spain | 2022 |
| EPI_ISL_19131538 | GISAID | RSV-A | Spain | 2022 |
| EPI_ISL_19131539 | GISAID | RSV-A | Spain | 2022 |
| EPI_ISL_19131541 | GISAID | RSV-A | Spain | 2022 |
| EPI_ISL_19131544 | GISAID | RSV-A | Spain | 2022 |
| EPI_ISL_19131547 | GISAID | RSV-A | Spain | 2022 |
| EPI_ISL_19131551 | GISAID | RSV-A | Spain | 2022 |
| EPI_ISL_19131552 | GISAID | RSV-A | Spain | 2022 |
| EPI_ISL_19131553 | GISAID | RSV-A | Spain | 2022 |
| EPI_ISL_19131555 | GISAID | RSV-A | Spain | 2022 |
| EPI_ISL_19131556 | GISAID | RSV-A | Spain | 2022 |
| EPI_ISL_19256949 | GISAID | RSV-A | Spain | 2024 |
| EPI_ISL_18682089 | GISAID | RSV-A | Spain | 2016 |
| EPI_ISL_18682093 | GISAID | RSV-A | Spain | 2018 |
| EPI_ISL_18682096 | GISAID | RSV-A | Spain | 2019 |
| EPI_ISL_18682098 | GISAID | RSV-A | Spain | 2021 |
| EPI_ISL_18682189 | GISAID | RSV-A | Spain | 2023 |

| | | | | |
|---|---|---|---|---|
| EPI_ISL_18682190 | GISAID | RSV-A | Spain | 2022 |
| EPI_ISL_18682193 | GISAID | RSV-A | Spain | 2023 |
| EPI_ISL_18682199 | GISAID | RSV-A | Spain | 2023 |
| EPI_ISL_19048774 | GISAID | RSV-A | Spain | 2019 |
| EPI_ISL_19131501 | GISAID | RSV-A | Spain | 2022 |
| EPI_ISL_19131506 | GISAID | RSV-A | Spain | 2022 |
| EPI_ISL_19131507 | GISAID | RSV-A | Spain | 2022 |
| EPI_ISL_19131511 | GISAID | RSV-A | Spain | 2022 |
| EPI_ISL_19131512 | GISAID | RSV-A | Spain | 2022 |
| EPI_ISL_19131514 | GISAID | RSV-A | Spain | 2022 |
| EPI_ISL_19131515 | GISAID | RSV-A | Spain | 2022 |
| EPI_ISL_19131516 | GISAID | RSV-A | Spain | 2022 |
| EPI_ISL_19131518 | GISAID | RSV-A | Spain | 2022 |
| EPI_ISL_19131601 | GISAID | RSV-A | Spain | 2023 |
| EPI_ISL_19131602 | GISAID | RSV-A | Spain | 2023 |
| EPI_ISL_19131604 | GISAID | RSV-A | Spain | 2023 |
| EPI_ISL_19131612 | GISAID | RSV-A | Spain | 2023 |
| EPI_ISL_19131615 | GISAID | RSV-A | Spain | 2023 |
| EPI_ISL_19131616 | GISAID | RSV-A | Spain | 2023 |
| EPI_ISL_19131619 | GISAID | RSV-A | Spain | 2023 |
| EPI_ISL_19131627 | GISAID | RSV-A | Spain | 2023 |
| EPI_ISL_19131634 | GISAID | RSV-A | Spain | 2023 |
| EPI_ISL_19131635 | GISAID | RSV-A | Spain | 2023 |
| EPI_ISL_19131639 | GISAID | RSV-A | Spain | 2022 |
| EPI_ISL_19049007 | GISAID | RSV-A | Taiwan | 2019 |
| EPI_ISL_19048407 | GISAID | RSV-A | Taiwan | 2019 |
| EPI_ISL_19049005 | GISAID | RSV-A | United Kingdom | 2019 |
| EPI_ISL_19049262 | GISAID | RSV-A | United Kingdom | 2018 |
| EPI_ISL_18845913 | GISAID | RSV-A | United Kingdom | 2017 |
| EPI_ISL_19048953 | GISAID | RSV-A | United Kingdom | 2018 |
| EPI_ISL_19048321 | GISAID | RSV-A | United Kingdom | 2019 |

| | | | | |
|---|---|---|---|---|
| EPI_ISL_19048656 | GISAID | RSV-A | United Kingdom | 2018 |
| EPI_ISL_18636305 | GISAID | RSV-A | USA | 2015 |
| EPI_ISL_18636306 | GISAID | RSV-A | USA | 2011 |
| EPI_ISL_18938196 | GISAID | RSV-A | USA | 2023 |
| EPI_ISL_18938240 | GISAID | RSV-A | USA | 2023 |
| EPI_ISL_18938266 | GISAID | RSV-A | USA | 2023 |
| EPI_ISL_18938268 | GISAID | RSV-A | USA | 2023 |
| EPI_ISL_18938283 | GISAID | RSV-A | USA | 2023 |
| EPI_ISL_18938292 | GISAID | RSV-A | USA | 2023 |
| EPI_ISL_18938297 | GISAID | RSV-A | USA | 2023 |
| EPI_ISL_18939330 | GISAID | RSV-A | USA | 2022 |
| EPI_ISL_18939331 | GISAID | RSV-A | USA | 2022 |
| EPI_ISL_18939332 | GISAID | RSV-A | USA | 2022 |
| EPI_ISL_18939340 | GISAID | RSV-A | USA | 2022 |
| EPI_ISL_18939344 | GISAID | RSV-A | USA | 2022 |
| EPI_ISL_18939366 | GISAID | RSV-A | USA | 2022 |
| EPI_ISL_18939368 | GISAID | RSV-A | USA | 2022 |
| EPI_ISL_18939381 | GISAID | RSV-A | USA | 2019 |
| EPI_ISL_18939384 | GISAID | RSV-A | USA | 2022 |
| EPI_ISL_18939387 | GISAID | RSV-A | USA | 2019 |
| EPI_ISL_18939388 | GISAID | RSV-A | USA | 2019 |
| EPI_ISL_18939397 | GISAID | RSV-A | USA | 2022 |
| EPI_ISL_18939398 | GISAID | RSV-A | USA | 2019 |
| EPI_ISL_18939400 | GISAID | RSV-A | USA | 2020 |
| EPI_ISL_18939406 | GISAID | RSV-A | USA | 2019 |
| EPI_ISL_18939409 | GISAID | RSV-A | USA | 2019 |
| EPI_ISL_18939421 | GISAID | RSV-A | USA | 2022 |
| EPI_ISL_18939422 | GISAID | RSV-A | USA | 2022 |
| EPI_ISL_18939453 | GISAID | RSV-A | USA | 2022 |
| EPI_ISL_18939459 | GISAID | RSV-A | USA | 2019 |
| EPI_ISL_18939472 | GISAID | RSV-A | USA | 2022 |
| EPI_ISL_18939473 | GISAID | RSV-A | USA | 2022 |

| | | | | |
|---|---|---|---|---|
| EPI_ISL_18939485 | GISAID | RSV-A | USA | 2022 |
| EPI_ISL_18939497 | GISAID | RSV-A | USA | 2020 |
| EPI_ISL_18939499 | GISAID | RSV-A | USA | 2018 |
| EPI_ISL_18939511 | GISAID | RSV-A | USA | 2022 |
| EPI_ISL_18939516 | GISAID | RSV-A | USA | 2022 |
| EPI_ISL_18939517 | GISAID | RSV-A | USA | 2022 |
| EPI_ISL_18939523 | GISAID | RSV-A | USA | 2019 |
| EPI_ISL_18939532 | GISAID | RSV-A | USA | 2022 |
| EPI_ISL_18939539 | GISAID | RSV-A | USA | 2022 |
| EPI_ISL_18939541 | GISAID | RSV-A | USA | 2018 |
| EPI_ISL_18939546 | GISAID | RSV-A | USA | 2022 |
| EPI_ISL_18939586 | GISAID | RSV-A | USA | 2024 |
| EPI_ISL_18939587 | GISAID | RSV-A | USA | 2024 |
| EPI_ISL_18939589 | GISAID | RSV-A | USA | 2024 |
| EPI_ISL_18939593 | GISAID | RSV-A | USA | 2024 |
| EPI_ISL_18939595 | GISAID | RSV-A | USA | 2024 |
| EPI_ISL_18939596 | GISAID | RSV-A | USA | 2024 |
| EPI_ISL_18939597 | GISAID | RSV-A | USA | 2024 |
| EPI_ISL_18939598 | GISAID | RSV-A | USA | 2024 |
| EPI_ISL_18939599 | GISAID | RSV-A | USA | 2024 |
| EPI_ISL_19006862 | GISAID | RSV-A | USA | 2020 |
| EPI_ISL_19006867 | GISAID | RSV-A | USA | 2019 |
| EPI_ISL_19006872 | GISAID | RSV-A | USA | 2019 |
| EPI_ISL_19006981 | GISAID | RSV-A | USA | 2020 |
| EPI_ISL_19006983 | GISAID | RSV-A | USA | 2020 |
| EPI_ISL_19006994 | GISAID | RSV-A | USA | 2020 |
| EPI_ISL_19007004 | GISAID | RSV-A | USA | 2019 |
| EPI_ISL_19026239 | GISAID | RSV-A | USA | 2023 |
| EPI_ISL_19026374 | GISAID | RSV-A | USA | 2015 |
| EPI_ISL_19060156 | GISAID | RSV-A | USA | 2024 |
| EPI_ISL_19069900 | GISAID | RSV-A | USA | 2022 |
| EPI_ISL_19069905 | GISAID | RSV-A | USA | 2022 |

| | | | | |
|---|---|---|---|---|
| EPI_ISL_19069908 | GISAID | RSV-A | USA | 2022 |
| EPI_ISL_19069914 | GISAID | RSV-A | USA | 2022 |
| EPI_ISL_19069916 | GISAID | RSV-A | USA | 2022 |
| EPI_ISL_19137618 | GISAID | RSV-A | USA | 2022 |
| EPI_ISL_19137627 | GISAID | RSV-A | USA | 2022 |
| EPI_ISL_19140041 | GISAID | RSV-A | USA | 2024 |
| EPI_ISL_19140056 | GISAID | RSV-A | USA | 2024 |
| EPI_ISL_18452368 | GISAID | RSV-A | USA | 2022 |
| EPI_ISL_18452369 | GISAID | RSV-A | USA | 2022 |
| EPI_ISL_18742322 | GISAID | RSV-A | USA | 2023 |
| EPI_ISL_18923253 | GISAID | RSV-A | USA | 2023 |
| EPI_ISL_18923258 | GISAID | RSV-A | USA | 2023 |
| EPI_ISL_18923283 | GISAID | RSV-A | USA | 2023 |
| EPI_ISL_18939603 | GISAID | RSV-A | USA | 2024 |
| EPI_ISL_18939609 | GISAID | RSV-A | USA | 2024 |
| EPI_ISL_18939611 | GISAID | RSV-A | USA | 2024 |
| EPI_ISL_19006822 | GISAID | RSV-A | USA | 2019 |
| EPI_ISL_19006823 | GISAID | RSV-A | USA | 2019 |
| EPI_ISL_19006826 | GISAID | RSV-A | USA | 2019 |
| EPI_ISL_19006829 | GISAID | RSV-A | USA | 2019 |
| EPI_ISL_19006845 | GISAID | RSV-A | USA | 2020 |
| EPI_ISL_19006849 | GISAID | RSV-A | USA | 2020 |
| EPI_ISL_19006854 | GISAID | RSV-A | USA | 2019 |
| EPI_ISL_19006973 | GISAID | RSV-A | USA | 2024 |
| EPI_ISL_19006974 | GISAID | RSV-A | USA | 2019 |
| EPI_ISL_19026306 | GISAID | RSV-A | USA | 2016 |
| EPI_ISL_19026310 | GISAID | RSV-A | USA | 2016 |
| EPI_ISL_19026311 | GISAID | RSV-A | USA | 2016 |
| EPI_ISL_19026312 | GISAID | RSV-A | USA | 2015 |
| EPI_ISL_19026316 | GISAID | RSV-A | USA | 2015 |
| EPI_ISL_19026320 | GISAID | RSV-A | USA | 2015 |
| EPI_ISL_19026322 | GISAID | RSV-A | USA | 2016 |

| | | | | |
|---|---|---|---|---|
| EPI_ISL_19026323 | GISAID | RSV-A | USA | 2016 |
| EPI_ISL_19026324 | GISAID | RSV-A | USA | 2017 |
| EPI_ISL_19069878 | GISAID | RSV-A | USA | 2022 |
| EPI_ISL_19069879 | GISAID | RSV-A | USA | 2022 |
| EPI_ISL_19069885 | GISAID | RSV-A | USA | 2022 |
| EPI_ISL_19069886 | GISAID | RSV-A | USA | 2022 |
| EPI_ISL_19069888 | GISAID | RSV-A | USA | 2022 |
| EPI_ISL_19231335 | GISAID | RSV-A | USA | 2024 |
| EPI_ISL_19231362 | GISAID | RSV-A | USA | 2024 |
| EPI_ISL_19231363 | GISAID | RSV-A | USA | 2024 |
| EPI_ISL_19231364 | GISAID | RSV-A | USA | 2024 |
| EPI_ISL_19231372 | GISAID | RSV-A | USA | 2024 |
| EPI_ISL_19237917 | GISAID | RSV-A | USA | 2024 |
| EPI_ISL_19237918 | GISAID | RSV-A | USA | 2024 |
| EPI_ISL_2588378 | GISAID | RSV-A | USA | 2013 |
| EPI_ISL_19006903 | GISAID | RSV-A | USA | 2019 |
| EPI_ISL_19006905 | GISAID | RSV-A | USA | 2019 |
| EPI_ISL_19006906 | GISAID | RSV-A | USA | 2020 |
| EPI_ISL_19006917 | GISAID | RSV-A | USA | 2019 |
| EPI_ISL_19006928 | GISAID | RSV-A | USA | 2020 |
| EPI_ISL_19069880 | GISAID | RSV-A | USA | 2023 |
| EPI_ISL_19069881 | GISAID | RSV-A | USA | 2022 |
| EPI_ISL_19069882 | GISAID | RSV-A | USA | 2022 |
| EPI_ISL_19069883 | GISAID | RSV-A | USA | 2022 |
| EPI_ISL_19069895 | GISAID | RSV-A | USA | 2022 |
| EPI_ISL_19231360 | GISAID | RSV-A | USA | 2024 |
| EPI_ISL_19231366 | GISAID | RSV-A | USA | 2024 |
| EPI_ISL_11055794 | GISAID | RSV-B | South Africa | 2021 |
| EPI_ISL_11055802 | GISAID | RSV-B | South Africa | 2021 |
| EPI_ISL_12970406 | GISAID | RSV-B | Philippines | 2019 |
| EPI_ISL_15055330 | GISAID | RSV-B | Argentina | 2018 |
| EPI_ISL_15055336 | GISAID | RSV-B | Argentina | 2018 |

| | | | | |
|---|---|---|---|---|
| EPI_ISL_15055345 | GISAID | RSV-B | Argentina | 2018 |
| EPI_ISL_15067695 | GISAID | RSV-B | Argentina | 2018 |
| EPI_ISL_15067701 | GISAID | RSV-B | Argentina | 2018 |
| EPI_ISL_15067710 | GISAID | RSV-B | Argentina | 2018 |
| EPI_ISL_15120685 | GISAID | RSV-B | Morocco | 2015 |
| EPI_ISL_15120686 | GISAID | RSV-B | Morocco | 2015 |
| EPI_ISL_15120691 | GISAID | RSV-B | Morocco | 2016 |
| EPI_ISL_15120693 | GISAID | RSV-B | Morocco | 2016 |
| EPI_ISL_15120695 | GISAID | RSV-B | Morocco | 2016 |
| EPI_ISL_15120696 | GISAID | RSV-B | Morocco | 2016 |
| EPI_ISL_15120697 | GISAID | RSV-B | Morocco | 2016 |
| EPI_ISL_15120698 | GISAID | RSV-B | Morocco | 2016 |
| EPI_ISL_15120699 | GISAID | RSV-B | Morocco | 2016 |
| EPI_ISL_15120700 | GISAID | RSV-B | Morocco | 2016 |
| EPI_ISL_15120701 | GISAID | RSV-B | Morocco | 2016 |
| EPI_ISL_15120702 | GISAID | RSV-B | Morocco | 2016 |
| EPI_ISL_15120704 | GISAID | RSV-B | Morocco | 2016 |
| EPI_ISL_15120707 | GISAID | RSV-B | Morocco | 2016 |
| EPI_ISL_15120708 | GISAID | RSV-B | Morocco | 2016 |
| EPI_ISL_15120709 | GISAID | RSV-B | Morocco | 2016 |
| EPI_ISL_15120710 | GISAID | RSV-B | Morocco | 2016 |
| EPI_ISL_15120711 | GISAID | RSV-B | Morocco | 2016 |
| EPI_ISL_15120712 | GISAID | RSV-B | Morocco | 2016 |
| EPI_ISL_15120714 | GISAID | RSV-B | Morocco | 2016 |
| EPI_ISL_15120722 | GISAID | RSV-B | Morocco | 2016 |
| EPI_ISL_15120724 | GISAID | RSV-B | Morocco | 2016 |
| EPI_ISL_15120725 | GISAID | RSV-B | Morocco | 2016 |
| EPI_ISL_15120733 | GISAID | RSV-B | Morocco | 2016 |
| EPI_ISL_15120734 | GISAID | RSV-B | Morocco | 2016 |
| EPI_ISL_15120735 | GISAID | RSV-B | Morocco | 2016 |
| EPI_ISL_15120737 | GISAID | RSV-B | Morocco | 2017 |
| EPI_ISL_15120740 | GISAID | RSV-B | Morocco | 2017 |

| | | | | |
|---|---|---|---|---|
| EPI_ISL_15120741 | GISAID | RSV-B | Morocco | 2017 |
| EPI_ISL_15120742 | GISAID | RSV-B | Morocco | 2017 |
| EPI_ISL_15120744 | GISAID | RSV-B | Morocco | 2017 |
| EPI_ISL_15120746 | GISAID | RSV-B | Morocco | 2017 |
| EPI_ISL_15120747 | GISAID | RSV-B | Morocco | 2017 |
| EPI_ISL_15120750 | GISAID | RSV-B | Morocco | 2017 |
| EPI_ISL_15120751 | GISAID | RSV-B | Morocco | 2017 |
| EPI_ISL_15120752 | GISAID | RSV-B | Morocco | 2017 |
| EPI_ISL_15120753 | GISAID | RSV-B | Morocco | 2017 |
| EPI_ISL_15120754 | GISAID | RSV-B | Morocco | 2017 |
| EPI_ISL_15120755 | GISAID | RSV-B | Morocco | 2017 |
| EPI_ISL_15120756 | GISAID | RSV-B | Morocco | 2017 |
| EPI_ISL_15120759 | GISAID | RSV-B | Morocco | 2017 |
| EPI_ISL_15120760 | GISAID | RSV-B | Morocco | 2017 |
| EPI_ISL_15120761 | GISAID | RSV-B | Morocco | 2017 |
| EPI_ISL_15120763 | GISAID | RSV-B | Morocco | 2017 |
| EPI_ISL_15120764 | GISAID | RSV-B | Morocco | 2017 |
| EPI_ISL_15120765 | GISAID | RSV-B | Morocco | 2017 |
| EPI_ISL_15120766 | GISAID | RSV-B | Morocco | 2017 |
| EPI_ISL_15120767 | GISAID | RSV-B | Morocco | 2017 |
| EPI_ISL_15120768 | GISAID | RSV-B | Morocco | 2017 |
| EPI_ISL_15120769 | GISAID | RSV-B | Morocco | 2017 |
| EPI_ISL_15120779 | GISAID | RSV-B | Morocco | 2017 |
| EPI_ISL_15120784 | GISAID | RSV-B | Morocco | 2018 |
| EPI_ISL_15120787 | GISAID | RSV-B | Morocco | 2019 |
| EPI_ISL_15120788 | GISAID | RSV-B | Morocco | 2019 |
| EPI_ISL_15120789 | GISAID | RSV-B | Morocco | 2019 |
| EPI_ISL_15120790 | GISAID | RSV-B | Morocco | 2019 |
| EPI_ISL_15120791 | GISAID | RSV-B | Morocco | 2019 |
| EPI_ISL_15120792 | GISAID | RSV-B | Morocco | 2019 |
| EPI_ISL_15120793 | GISAID | RSV-B | Morocco | 2019 |
| EPI_ISL_15120794 | GISAID | RSV-B | Morocco | 2019 |

| | | | | |
|---|---|---|---|---|
| EPI_ISL_15120796 | GISAID | RSV-B | Morocco | 2019 |
| EPI_ISL_15120797 | GISAID | RSV-B | Morocco | 2019 |
| EPI_ISL_15120798 | GISAID | RSV-B | Morocco | 2019 |
| EPI_ISL_15120799 | GISAID | RSV-B | Morocco | 2019 |
| EPI_ISL_15752977 | GISAID | RSV-B | Spain | 2019 |
| EPI_ISL_15752979 | GISAID | RSV-B | Spain | 2019 |
| EPI_ISL_15752996 | GISAID | RSV-B | United Kingdom | 2019 |
| EPI_ISL_15753016 | GISAID | RSV-B | Spain | 2019 |
| EPI_ISL_15753041 | GISAID | RSV-B | United Kingdom | 2018 |
| EPI_ISL_15753050 | GISAID | RSV-B | Spain | 2020 |
| EPI_ISL_15753087 | GISAID | RSV-B | Spain | 2020 |
| EPI_ISL_15753092 | GISAID | RSV-B | Spain | 2019 |
| EPI_ISL_15753103 | GISAID | RSV-B | United Kingdom | 2019 |
| EPI_ISL_15753116 | GISAID | RSV-B | United Kingdom | 2018 |
| EPI_ISL_15753119 | GISAID | RSV-B | United Kingdom | 2018 |
| EPI_ISL_15753133 | GISAID | RSV-B | United Kingdom | 2019 |
| EPI_ISL_15753134 | GISAID | RSV-B | Spain | 2020 |
| EPI_ISL_15753148 | GISAID | RSV-B | United Kingdom | 2018 |
| EPI_ISL_15753164 | GISAID | RSV-B | Spain | 2019 |
| EPI_ISL_15753172 | GISAID | RSV-B | Spain | 2020 |
| EPI_ISL_15753202 | GISAID | RSV-B | Netherlands | 2018 |
| EPI_ISL_1647506 | GISAID | RSV-B | England | 2019 |
| EPI_ISL_1647509 | GISAID | RSV-B | England | 2019 |
| EPI_ISL_1647511 | GISAID | RSV-B | England | 2019 |
| EPI_ISL_1647517 | GISAID | RSV-B | England | 2019 |
| EPI_ISL_1647523 | GISAID | RSV-B | England | 2019 |
| EPI_ISL_1647546 | GISAID | RSV-B | England | 2019 |
| EPI_ISL_1647555 | GISAID | RSV-B | England | 2019 |
| EPI_ISL_1647575 | GISAID | RSV-B | England | 2020 |
| EPI_ISL_1647579 | GISAID | RSV-B | England | 2020 |

| | | | | |
|---|---|---|---|---|
| EPI_ISL_1647583 | GISAID | RSV-B | England | 2020 |
| EPI_ISL_1647585 | GISAID | RSV-B | England | 2020 |
| EPI_ISL_1647599 | GISAID | RSV-B | England | 2020 |
| EPI_ISL_1653996 | GISAID | RSV-B | Australia | 2019 |
| EPI_ISL_1653999 | GISAID | RSV-B | Australia | 2019 |
| EPI_ISL_16709010 | GISAID | RSV-B | England | 2021 |
| EPI_ISL_16714336 | GISAID | RSV-B | England | 2021 |
| EPI_ISL_16714389 | GISAID | RSV-B | England | 2021 |
| EPI_ISL_16714576 | GISAID | RSV-B | England | 2022 |
| EPI_ISL_16714657 | GISAID | RSV-B | England | 2021 |
| EPI_ISL_17089188 | GISAID | RSV-B | USA | 2019 |
| EPI_ISL_17221668 | GISAID | RSV-B | USA | 2019 |
| EPI_ISL_17417590 | GISAID | RSV-B | Philippines | 2019 |
| EPI_ISL_1760385 | GISAID | RSV-B | Australia | 2018 |
| EPI_ISL_1760399 | GISAID | RSV-B | Australia | 2018 |
| EPI_ISL_1760400 | GISAID | RSV-B | Australia | 2017 |
| EPI_ISL_1760402 | GISAID | RSV-B | Australia | 2017 |
| EPI_ISL_1760438 | GISAID | RSV-B | Australia | 2019 |
| EPI_ISL_1760439 | GISAID | RSV-B | Australia | 2018 |
| EPI_ISL_1760441 | GISAID | RSV-B | Australia | 2018 |
| EPI_ISL_1760442 | GISAID | RSV-B | Australia | 2018 |
| EPI_ISL_1760443 | GISAID | RSV-B | Australia | 2019 |
| EPI_ISL_1760444 | GISAID | RSV-B | Australia | 2018 |
| EPI_ISL_18005802 | GISAID | RSV-B | Germany | 2019 |
| EPI_ISL_18005803 | GISAID | RSV-B | Germany | 2019 |
| EPI_ISL_18005804 | GISAID | RSV-B | Germany | 2019 |
| EPI_ISL_18090576 | GISAID | RSV-B | Australia | 2018 |
| EPI_ISL_18090584 | GISAID | RSV-B | Australia | 2018 |
| EPI_ISL_18090592 | GISAID | RSV-B | Australia | 2018 |
| EPI_ISL_18090672 | GISAID | RSV-B | Australia | 2018 |
| EPI_ISL_18143574 | GISAID | RSV-B | USA | 2020 |
| EPI_ISL_18143697 | GISAID | RSV-B | USA | 2019 |

| | | | | |
|---|---|---|---|---|
| EPI_ISL_18277124 | GISAID | RSV-B | Scotland | 2021 |
| EPI_ISL_18321026 | GISAID | RSV-B | Ireland | 2022 |
| EPI_ISL_18321028 | GISAID | RSV-B | Ireland | 2022 |
| EPI_ISL_18321030 | GISAID | RSV-B | Ireland | 2022 |
| EPI_ISL_18321032 | GISAID | RSV-B | Ireland | 2022 |
| EPI_ISL_18321057 | GISAID | RSV-B | Ireland | 2022 |
| EPI_ISL_18321065 | GISAID | RSV-B | Ireland | 2022 |
| EPI_ISL_18321067 | GISAID | RSV-B | Ireland | 2022 |
| EPI_ISL_18321070 | GISAID | RSV-B | Ireland | 2022 |
| EPI_ISL_18321074 | GISAID | RSV-B | Ireland | 2022 |
| EPI_ISL_18321077 | GISAID | RSV-B | Ireland | 2022 |
| EPI_ISL_18321105 | GISAID | RSV-B | Ireland | 2022 |
| EPI_ISL_18321107 | GISAID | RSV-B | Ireland | 2022 |
| EPI_ISL_18321110 | GISAID | RSV-B | Ireland | 2022 |
| EPI_ISL_18321112 | GISAID | RSV-B | Ireland | 2022 |
| EPI_ISL_18321114 | GISAID | RSV-B | Ireland | 2022 |
| EPI_ISL_18321118 | GISAID | RSV-B | Ireland | 2022 |
| EPI_ISL_18321119 | GISAID | RSV-B | Ireland | 2022 |
| EPI_ISL_18321120 | GISAID | RSV-B | Ireland | 2022 |
| EPI_ISL_18321122 | GISAID | RSV-B | Ireland | 2022 |
| EPI_ISL_18321123 | GISAID | RSV-B | Ireland | 2022 |
| EPI_ISL_18321129 | GISAID | RSV-B | Ireland | 2022 |
| EPI_ISL_18321132 | GISAID | RSV-B | Ireland | 2022 |
| EPI_ISL_18321133 | GISAID | RSV-B | Ireland | 2022 |
| EPI_ISL_18321135 | GISAID | RSV-B | Ireland | 2022 |
| EPI_ISL_18321137 | GISAID | RSV-B | Ireland | 2022 |
| EPI_ISL_18321139 | GISAID | RSV-B | Ireland | 2022 |
| EPI_ISL_18321140 | GISAID | RSV-B | Ireland | 2022 |
| EPI_ISL_18321150 | GISAID | RSV-B | Ireland | 2022 |
| EPI_ISL_18321154 | GISAID | RSV-B | Ireland | 2022 |
| EPI_ISL_18321155 | GISAID | RSV-B | Ireland | 2022 |
| EPI_ISL_18321158 | GISAID | RSV-B | Ireland | 2022 |

| | | | | |
|---|---|---|---|---|
| EPI_ISL_1834176 | GISAID | RSV-B | England | 2019 |
| EPI_ISL_1834182 | GISAID | RSV-B | England | 2020 |
| EPI_ISL_18482806 | GISAID | RSV-B | China | 2019 |
| EPI_ISL_18591794 | GISAID | RSV-B | England | 2023 |
| EPI_ISL_18591810 | GISAID | RSV-B | England | 2023 |
| EPI_ISL_18591811 | GISAID | RSV-B | England | 2023 |
| EPI_ISL_18591813 | GISAID | RSV-B | England | 2023 |
| EPI_ISL_18622389 | GISAID | RSV-B | Brazil | 2023 |
| EPI_ISL_18622390 | GISAID | RSV-B | Brazil | 2023 |
| EPI_ISL_18622402 | GISAID | RSV-B | Brazil | 2023 |
| EPI_ISL_18622404 | GISAID | RSV-B | Brazil | 2023 |
| EPI_ISL_18648348 | GISAID | RSV-B | Spain | 2023 |
| EPI_ISL_18648350 | GISAID | RSV-B | Spain | 2023 |
| EPI_ISL_18653611 | GISAID | RSV-B | Australia | 2023 |
| EPI_ISL_18656998 | GISAID | RSV-B | Australia | 2023 |
| EPI_ISL_18656999 | GISAID | RSV-B | Australia | 2023 |
| EPI_ISL_18657000 | GISAID | RSV-B | Australia | 2023 |
| EPI_ISL_18657010 | GISAID | RSV-B | Australia | 2023 |
| EPI_ISL_18657021 | GISAID | RSV-B | Australia | 2023 |
| EPI_ISL_18657024 | GISAID | RSV-B | Australia | 2023 |
| EPI_ISL_18657027 | GISAID | RSV-B | Australia | 2023 |
| EPI_ISL_18657028 | GISAID | RSV-B | Australia | 2023 |
| EPI_ISL_18657029 | GISAID | RSV-B | Australia | 2023 |
| EPI_ISL_18657032 | GISAID | RSV-B | Australia | 2023 |
| EPI_ISL_18657033 | GISAID | RSV-B | Australia | 2023 |
| EPI_ISL_18657040 | GISAID | RSV-B | Australia | 2023 |
| EPI_ISL_18657054 | GISAID | RSV-B | Australia | 2023 |
| EPI_ISL_18657059 | GISAID | RSV-B | Australia | 2023 |
| EPI_ISL_18682099 | GISAID | RSV-B | Spain | 2021 |
| EPI_ISL_18682102 | GISAID | RSV-B | Spain | 2021 |
| EPI_ISL_18682106 | GISAID | RSV-B | Spain | 2021 |
| EPI_ISL_18682108 | GISAID | RSV-B | Spain | 2021 |

| | | | | |
|---|---|---|---|---|
| EPI_ISL_18682110 | GISAID | RSV-B | Spain | 2021 |
| EPI_ISL_18682112 | GISAID | RSV-B | Spain | 2021 |
| EPI_ISL_18682113 | GISAID | RSV-B | Spain | 2021 |
| EPI_ISL_18682116 | GISAID | RSV-B | Spain | 2021 |
| EPI_ISL_18682121 | GISAID | RSV-B | Spain | 2021 |
| EPI_ISL_18682123 | GISAID | RSV-B | Spain | 2022 |
| EPI_ISL_18682126 | GISAID | RSV-B | Spain | 2022 |
| EPI_ISL_18682136 | GISAID | RSV-B | Spain | 2021 |
| EPI_ISL_18682138 | GISAID | RSV-B | Spain | 2021 |
| EPI_ISL_18682139 | GISAID | RSV-B | Spain | 2021 |
| EPI_ISL_18682140 | GISAID | RSV-B | Spain | 2021 |
| EPI_ISL_18682144 | GISAID | RSV-B | Spain | 2021 |
| EPI_ISL_18682148 | GISAID | RSV-B | Spain | 2021 |
| EPI_ISL_18682149 | GISAID | RSV-B | Spain | 2021 |
| EPI_ISL_18682154 | GISAID | RSV-B | Spain | 2022 |
| EPI_ISL_18682156 | GISAID | RSV-B | Spain | 2022 |
| EPI_ISL_18682160 | GISAID | RSV-B | Spain | 2021 |
| EPI_ISL_18682163 | GISAID | RSV-B | Spain | 2021 |
| EPI_ISL_18682165 | GISAID | RSV-B | Spain | 2021 |
| EPI_ISL_18682174 | GISAID | RSV-B | Spain | 2022 |
| EPI_ISL_18682176 | GISAID | RSV-B | Spain | 2021 |
| EPI_ISL_18682179 | GISAID | RSV-B | Spain | 2022 |
| EPI_ISL_18682182 | GISAID | RSV-B | Spain | 2023 |
| EPI_ISL_18682185 | GISAID | RSV-B | Spain | 2022 |
| EPI_ISL_18682188 | GISAID | RSV-B | Spain | 2022 |
| EPI_ISL_18682192 | GISAID | RSV-B | Spain | 2023 |
| EPI_ISL_18682194 | GISAID | RSV-B | Spain | 2021 |
| EPI_ISL_18682196 | GISAID | RSV-B | Spain | 2022 |
| EPI_ISL_18682202 | GISAID | RSV-B | Spain | 2022 |
| EPI_ISL_18682203 | GISAID | RSV-B | Spain | 2022 |
| EPI_ISL_18682207 | GISAID | RSV-B | Spain | 2021 |
| EPI_ISL_18682208 | GISAID | RSV-B | Spain | 2022 |

| | | | | |
|---|---|---|---|---|
| EPI_ISL_18682217 | GISAID | RSV-B | Spain | 2023 |
| EPI_ISL_18682218 | GISAID | RSV-B | Spain | 2023 |
| EPI_ISL_18682239 | GISAID | RSV-B | Spain | 2018 |
| EPI_ISL_18682252 | GISAID | RSV-B | Spain | 2020 |
| EPI_ISL_18682254 | GISAID | RSV-B | Spain | 2021 |
| EPI_ISL_18682277 | GISAID | RSV-B | Spain | 2016 |
| EPI_ISL_18682291 | GISAID | RSV-B | Spain | 2018 |
| EPI_ISL_18682312 | GISAID | RSV-B | Spain | 2021 |
| EPI_ISL_18682330 | GISAID | RSV-B | Spain | 2018 |
| EPI_ISL_18682331 | GISAID | RSV-B | Spain | 2018 |
| EPI_ISL_18682339 | GISAID | RSV-B | Spain | 2021 |
| EPI_ISL_18682341 | GISAID | RSV-B | Spain | 2021 |
| EPI_ISL_18682371 | GISAID | RSV-B | Spain | 2017 |
| EPI_ISL_18682377 | GISAID | RSV-B | Spain | 2021 |
| EPI_ISL_18682396 | GISAID | RSV-B | Spain | 2017 |
| EPI_ISL_18682398 | GISAID | RSV-B | Spain | 2018 |
| EPI_ISL_18682402 | GISAID | RSV-B | Spain | 2021 |
| EPI_ISL_18682409 | GISAID | RSV-B | Spain | 2018 |
| EPI_ISL_18682410 | GISAID | RSV-B | Spain | 2018 |
| EPI_ISL_18682422 | GISAID | RSV-B | Spain | 2019 |
| EPI_ISL_18682424 | GISAID | RSV-B | Spain | 2020 |
| EPI_ISL_18682442 | GISAID | RSV-B | Spain | 2020 |
| EPI_ISL_18682444 | GISAID | RSV-B | Spain | 2018 |
| EPI_ISL_18694963 | GISAID | RSV-B | USA | 2023 |
| EPI_ISL_18694965 | GISAID | RSV-B | USA | 2023 |
| EPI_ISL_18694974 | GISAID | RSV-B | USA | 2023 |
| EPI_ISL_18694987 | GISAID | RSV-B | USA | 2021 |
| EPI_ISL_18695001 | GISAID | RSV-B | USA | 2023 |
| EPI_ISL_18695032 | GISAID | RSV-B | USA | 2023 |
| EPI_ISL_18695035 | GISAID | RSV-B | USA | 2021 |
| EPI_ISL_18695055 | GISAID | RSV-B | USA | 2022 |
| EPI_ISL_18695057 | GISAID | RSV-B | USA | 2021 |

| | | | | |
|---|---|---|---|---|
| EPI_ISL_18695060 | GISAID | RSV-B | USA | 2023 |
| EPI_ISL_18695063 | GISAID | RSV-B | USA | 2023 |
| EPI_ISL_18698533 | GISAID | RSV-B | Canary Islands | 2023 |
| EPI_ISL_18698544 | GISAID | RSV-B | Canary Islands | 2023 |
| EPI_ISL_18698590 | GISAID | RSV-B | Canary Islands | 2022 |
| EPI_ISL_18698591 | GISAID | RSV-B | Canary Islands | 2022 |
| EPI_ISL_18698598 | GISAID | RSV-B | Canary Islands | 2022 |
| EPI_ISL_18698626 | GISAID | RSV-B | Canary Islands | 2022 |
| EPI_ISL_18698629 | GISAID | RSV-B | Canary Islands | 2022 |
| EPI_ISL_18698642 | GISAID | RSV-B | Canary Islands | 2023 |
| EPI_ISL_18698666 | GISAID | RSV-B | Canary Islands | 2022 |
| EPI_ISL_18698677 | GISAID | RSV-B | Canary Islands | 2022 |
| EPI_ISL_18698679 | GISAID | RSV-B | Canary Islands | 2023 |
| EPI_ISL_18698700 | GISAID | RSV-B | Canary Islands | 2023 |
| EPI_ISL_18698706 | GISAID | RSV-B | Canary Islands | 2023 |
| EPI_ISL_18708128 | GISAID | RSV-B | South Africa | 2023 |
| EPI_ISL_18717756 | GISAID | RSV-B | England | 2023 |
| EPI_ISL_18717771 | GISAID | RSV-B | England | 2023 |
| EPI_ISL_18717774 | GISAID | RSV-B | England | 2023 |
| EPI_ISL_18725019 | GISAID | RSV-B | England | 2023 |
| EPI_ISL_18725032 | GISAID | RSV-B | England | 2023 |
| EPI_ISL_18725034 | GISAID | RSV-B | England | 2023 |
| EPI_ISL_18725037 | GISAID | RSV-B | England | 2023 |
| EPI_ISL_18725049 | GISAID | RSV-B | England | 2023 |
| EPI_ISL_18725070 | GISAID | RSV-B | England | 2023 |
| EPI_ISL_18725088 | GISAID | RSV-B | England | 2023 |
| EPI_ISL_18725100 | GISAID | RSV-B | England | 2023 |
| EPI_ISL_18725108 | GISAID | RSV-B | England | 2023 |
| EPI_ISL_18742340 | GISAID | RSV-B | USA | 2023 |
| EPI_ISL_18789017 | GISAID | RSV-B | France | 2019 |
| EPI_ISL_18789130 | GISAID | RSV-B | France | 2020 |
| EPI_ISL_18810653 | GISAID | RSV-B | Canary Islands | 2022 |

| | | | | |
|---|---|---|---|---|
| EPI_ISL_18810659 | GISAID | RSV-B | Canary Islands | 2022 |
| EPI_ISL_18810662 | GISAID | RSV-B | Canary Islands | 2022 |
| EPI_ISL_18810663 | GISAID | RSV-B | Canary Islands | 2022 |
| EPI_ISL_18810664 | GISAID | RSV-B | Canary Islands | 2022 |
| EPI_ISL_18810675 | GISAID | RSV-B | Canary Islands | 2022 |
| EPI_ISL_18810676 | GISAID | RSV-B | Canary Islands | 2022 |
| EPI_ISL_18810678 | GISAID | RSV-B | Canary Islands | 2022 |
| EPI_ISL_18810679 | GISAID | RSV-B | Canary Islands | 2022 |
| EPI_ISL_18810681 | GISAID | RSV-B | Canary Islands | 2022 |
| EPI_ISL_18810697 | GISAID | RSV-B | Canary Islands | 2023 |
| EPI_ISL_18810705 | GISAID | RSV-B | Canary Islands | 2023 |
| EPI_ISL_18846466 | GISAID | RSV-B | Spain | 2024 |
| EPI_ISL_18846474 | GISAID | RSV-B | Spain | 2024 |
| EPI_ISL_18919678 | GISAID | RSV-B | USA | 2022 |
| EPI_ISL_18927331 | GISAID | RSV-B | Spain | 2024 |
| EPI_ISL_18930952 | GISAID | RSV-B | England | 2023 |
| EPI_ISL_18930960 | GISAID | RSV-B | England | 2023 |
| EPI_ISL_18930992 | GISAID | RSV-B | England | 2024 |
| EPI_ISL_18931000 | GISAID | RSV-B | England | 2024 |
| EPI_ISL_18931002 | GISAID | RSV-B | England | 2024 |
| EPI_ISL_18931003 | GISAID | RSV-B | England | 2024 |
| EPI_ISL_18931015 | GISAID | RSV-B | England | 2024 |
| EPI_ISL_18931019 | GISAID | RSV-B | England | 2024 |
| EPI_ISL_18931609 | GISAID | RSV-B | Senegal | 2023 |
| EPI_ISL_18933022 | GISAID | RSV-B | Senegal | 2023 |
| EPI_ISL_18933030 | GISAID | RSV-B | Senegal | 2023 |
| EPI_ISL_18938210 | GISAID | RSV-B | USA | 2023 |
| EPI_ISL_18938211 | GISAID | RSV-B | USA | 2023 |
| EPI_ISL_18938221 | GISAID | RSV-B | USA | 2023 |
| EPI_ISL_18938252 | GISAID | RSV-B | USA | 2023 |
| EPI_ISL_18938265 | GISAID | RSV-B | USA | 2023 |
| EPI_ISL_18938277 | GISAID | RSV-B | USA | 2023 |

| | | | | |
|---|---|---|---|---|
| EPI_ISL_18938280 | GISAID | RSV-B | USA | 2023 |
| EPI_ISL_18938286 | GISAID | RSV-B | USA | 2023 |
| EPI_ISL_18938293 | GISAID | RSV-B | USA | 2023 |
| EPI_ISL_18938298 | GISAID | RSV-B | USA | 2023 |
| EPI_ISL_18939333 | GISAID | RSV-B | USA | 2021 |
| EPI_ISL_18939339 | GISAID | RSV-B | USA | 2018 |
| EPI_ISL_18939352 | GISAID | RSV-B | USA | 2022 |
| EPI_ISL_18939354 | GISAID | RSV-B | USA | 2018 |
| EPI_ISL_18939356 | GISAID | RSV-B | USA | 2020 |
| EPI_ISL_18939359 | GISAID | RSV-B | USA | 2021 |
| EPI_ISL_18939364 | GISAID | RSV-B | USA | 2019 |
| EPI_ISL_18939372 | GISAID | RSV-B | USA | 2018 |
| EPI_ISL_18939373 | GISAID | RSV-B | USA | 2018 |
| EPI_ISL_18939376 | GISAID | RSV-B | USA | 2018 |
| EPI_ISL_18939380 | GISAID | RSV-B | USA | 2019 |
| EPI_ISL_18939383 | GISAID | RSV-B | USA | 2019 |
| EPI_ISL_18939390 | GISAID | RSV-B | USA | 2018 |
| EPI_ISL_18939399 | GISAID | RSV-B | USA | 2021 |
| EPI_ISL_18939403 | GISAID | RSV-B | USA | 2021 |
| EPI_ISL_18939404 | GISAID | RSV-B | USA | 2021 |
| EPI_ISL_18939408 | GISAID | RSV-B | USA | 2021 |
| EPI_ISL_18939412 | GISAID | RSV-B | USA | 2021 |
| EPI_ISL_18939428 | GISAID | RSV-B | USA | 2018 |
| EPI_ISL_18939435 | GISAID | RSV-B | USA | 2022 |
| EPI_ISL_18939442 | GISAID | RSV-B | USA | 2021 |
| EPI_ISL_18939446 | GISAID | RSV-B | USA | 2021 |
| EPI_ISL_18939454 | GISAID | RSV-B | USA | 2021 |
| EPI_ISL_18939456 | GISAID | RSV-B | USA | 2020 |
| EPI_ISL_18939464 | GISAID | RSV-B | USA | 2021 |
| EPI_ISL_18939468 | GISAID | RSV-B | USA | 2018 |
| EPI_ISL_18939470 | GISAID | RSV-B | USA | 2021 |
| EPI_ISL_18939478 | GISAID | RSV-B | USA | 2018 |

| | | | | |
|---|---|---|---|---|
| EPI_ISL_18939482 | GISAID | RSV-B | USA | 2021 |
| EPI_ISL_18939483 | GISAID | RSV-B | USA | 2021 |
| EPI_ISL_18939492 | GISAID | RSV-B | USA | 2018 |
| EPI_ISL_18939493 | GISAID | RSV-B | USA | 2022 |
| EPI_ISL_18939498 | GISAID | RSV-B | USA | 2021 |
| EPI_ISL_18939507 | GISAID | RSV-B | USA | 2021 |
| EPI_ISL_18939508 | GISAID | RSV-B | USA | 2021 |
| EPI_ISL_18939519 | GISAID | RSV-B | USA | 2021 |
| EPI_ISL_18939529 | GISAID | RSV-B | USA | 2018 |
| EPI_ISL_18939530 | GISAID | RSV-B | USA | 2021 |
| EPI_ISL_18939531 | GISAID | RSV-B | USA | 2019 |
| EPI_ISL_18939535 | GISAID | RSV-B | USA | 2021 |
| EPI_ISL_18939536 | GISAID | RSV-B | USA | 2017 |
| EPI_ISL_18939538 | GISAID | RSV-B | USA | 2022 |
| EPI_ISL_18939545 | GISAID | RSV-B | USA | 2021 |
| EPI_ISL_18939582 | GISAID | RSV-B | USA | 2024 |
| EPI_ISL_18939585 | GISAID | RSV-B | USA | 2024 |
| EPI_ISL_18939594 | GISAID | RSV-B | USA | 2024 |
| EPI_ISL_18939607 | GISAID | RSV-B | USA | 2024 |
| EPI_ISL_18939610 | GISAID | RSV-B | USA | 2024 |
| EPI_ISL_18954891 | GISAID | RSV-B | Spain | 2023 |
| EPI_ISL_18954892 | GISAID | RSV-B | Spain | 2023 |
| EPI_ISL_18956304 | GISAID | RSV-B | USA | 2023 |
| EPI_ISL_18961700 | GISAID | RSV-B | Thailand | 2023 |
| EPI_ISL_18972338 | GISAID | RSV-B | Spain | 2024 |
| EPI_ISL_18972339 | GISAID | RSV-B | Spain | 2024 |
| EPI_ISL_18972340 | GISAID | RSV-B | Spain | 2024 |
| EPI_ISL_18972341 | GISAID | RSV-B | Spain | 2024 |
| EPI_ISL_18972342 | GISAID | RSV-B | Spain | 2024 |
| EPI_ISL_18972344 | GISAID | RSV-B | Spain | 2024 |
| EPI_ISL_18972345 | GISAID | RSV-B | Spain | 2024 |
| EPI_ISL_18972348 | GISAID | RSV-B | Spain | 2024 |

| | | | | |
|---|---|---|---|---|
| EPI_ISL_18972351 | GISAID | RSV-B | Spain | 2024 |
| EPI_ISL_18972352 | GISAID | RSV-B | Spain | 2024 |
| EPI_ISL_18972355 | GISAID | RSV-B | Spain | 2024 |
| EPI_ISL_18972358 | GISAID | RSV-B | Spain | 2024 |
| EPI_ISL_18972361 | GISAID | RSV-B | Spain | 2024 |
| EPI_ISL_18972362 | GISAID | RSV-B | Spain | 2024 |
| EPI_ISL_18972363 | GISAID | RSV-B | Spain | 2024 |
| EPI_ISL_18972364 | GISAID | RSV-B | Spain | 2024 |
| EPI_ISL_18972370 | GISAID | RSV-B | Spain | 2024 |
| EPI_ISL_18972371 | GISAID | RSV-B | Spain | 2024 |
| EPI_ISL_18973154 | GISAID | RSV-B | England | 2024 |
| EPI_ISL_18973156 | GISAID | RSV-B | England | 2024 |
| EPI_ISL_18973158 | GISAID | RSV-B | England | 2024 |
| EPI_ISL_18973162 | GISAID | RSV-B | England | 2024 |
| EPI_ISL_18980430 | GISAID | RSV-B | Spain | 2024 |
| EPI_ISL_18999163 | GISAID | RSV-B | Brazil | 2023 |
| EPI_ISL_19001893 | GISAID | RSV-B | Spain | 2023 |
| EPI_ISL_19001895 | GISAID | RSV-B | Spain | 2023 |
| EPI_ISL_19001896 | GISAID | RSV-B | Spain | 2023 |
| EPI_ISL_19001897 | GISAID | RSV-B | Spain | 2023 |
| EPI_ISL_19001898 | GISAID | RSV-B | Spain | 2023 |
| EPI_ISL_19001903 | GISAID | RSV-B | Spain | 2023 |
| EPI_ISL_19001904 | GISAID | RSV-B | Spain | 2023 |
| EPI_ISL_19001905 | GISAID | RSV-B | Spain | 2023 |
| EPI_ISL_19002032 | GISAID | RSV-B | Spain | 2024 |
| EPI_ISL_19002047 | GISAID | RSV-B | Spain | 2024 |
| EPI_ISL_19002049 | GISAID | RSV-B | Spain | 2024 |
| EPI_ISL_19002051 | GISAID | RSV-B | Spain | 2024 |
| EPI_ISL_19002067 | GISAID | RSV-B | Spain | 2023 |
| EPI_ISL_19002073 | GISAID | RSV-B | Spain | 2024 |
| EPI_ISL_19005575 | GISAID | RSV-B | Australia | 2023 |
| EPI_ISL_19005605 | GISAID | RSV-B | Australia | 2023 |

| | | | | |
|---|---|---|---|---|
| EPI_ISL_19005647 | GISAID | RSV-B | Australia | 2023 |
| EPI_ISL_19005650 | GISAID | RSV-B | Australia | 2023 |
| EPI_ISL_19005654 | GISAID | RSV-B | Australia | 2023 |
| EPI_ISL_19005658 | GISAID | RSV-B | Australia | 2023 |
| EPI_ISL_19005684 | GISAID | RSV-B | Australia | 2023 |
| EPI_ISL_19005688 | GISAID | RSV-B | Australia | 2023 |
| EPI_ISL_19005692 | GISAID | RSV-B | Australia | 2023 |
| EPI_ISL_19005698 | GISAID | RSV-B | Australia | 2023 |
| EPI_ISL_19005705 | GISAID | RSV-B | Australia | 2023 |
| EPI_ISL_19005706 | GISAID | RSV-B | Australia | 2023 |
| EPI_ISL_19005709 | GISAID | RSV-B | Australia | 2023 |
| EPI_ISL_19006831 | GISAID | RSV-B | USA | 2019 |
| EPI_ISL_19006833 | GISAID | RSV-B | USA | 2021 |
| EPI_ISL_19006865 | GISAID | RSV-B | USA | 2021 |
| EPI_ISL_19006866 | GISAID | RSV-B | USA | 2021 |
| EPI_ISL_19006877 | GISAID | RSV-B | USA | 2022 |
| EPI_ISL_19006887 | GISAID | RSV-B | USA | 2021 |
| EPI_ISL_19006898 | GISAID | RSV-B | USA | 2021 |
| EPI_ISL_19006900 | GISAID | RSV-B | USA | 2021 |
| EPI_ISL_19006909 | GISAID | RSV-B | USA | 2021 |
| EPI_ISL_19006920 | GISAID | RSV-B | USA | 2019 |
| EPI_ISL_19006924 | GISAID | RSV-B | USA | 2021 |
| EPI_ISL_19006929 | GISAID | RSV-B | USA | 2019 |
| EPI_ISL_19006932 | GISAID | RSV-B | USA | 2021 |
| EPI_ISL_19006936 | GISAID | RSV-B | USA | 2021 |
| EPI_ISL_19006955 | GISAID | RSV-B | USA | 2020 |
| EPI_ISL_19006961 | GISAID | RSV-B | USA | 2021 |
| EPI_ISL_19006977 | GISAID | RSV-B | USA | 2024 |
| EPI_ISL_19006978 | GISAID | RSV-B | USA | 2021 |
| EPI_ISL_19006988 | GISAID | RSV-B | USA | 2020 |
| EPI_ISL_19006993 | GISAID | RSV-B | USA | 2020 |
| EPI_ISL_19006996 | GISAID | RSV-B | USA | 2020 |

| | | | | |
|---|---|---|---|---|
| EPI_ISL_19007007 | GISAID | RSV-B | USA | 2022 |
| EPI_ISL_19014347 | GISAID | RSV-B | England | 2024 |
| EPI_ISL_19015689 | GISAID | RSV-B | Australia | 2023 |
| EPI_ISL_19015693 | GISAID | RSV-B | Australia | 2023 |
| EPI_ISL_19015775 | GISAID | RSV-B | Australia | 2023 |
| EPI_ISL_19026229 | GISAID | RSV-B | USA | 2024 |
| EPI_ISL_19026249 | GISAID | RSV-B | USA | 2023 |
| EPI_ISL_19026261 | GISAID | RSV-B | USA | 2023 |
| EPI_ISL_19026262 | GISAID | RSV-B | USA | 2024 |
| EPI_ISL_19028660 | GISAID | RSV-B | Italy | 2024 |
| EPI_ISL_19047957 | GISAID | RSV-B | Canada | 2018 |
| EPI_ISL_19047960 | GISAID | RSV-B | Germany | 2019 |
| EPI_ISL_19047981 | GISAID | RSV-B | Canada | 2018 |
| EPI_ISL_19048003 | GISAID | RSV-B | Australia | 2018 |
| EPI_ISL_19048037 | GISAID | RSV-B | Spain | 2020 |
| EPI_ISL_19048104 | GISAID | RSV-B | South Africa | 2018 |
| EPI_ISL_19048122 | GISAID | RSV-B | South Africa | 2019 |
| EPI_ISL_19048136 | GISAID | RSV-B | Finland | 2019 |
| EPI_ISL_19048154 | GISAID | RSV-B | Japan | 2018 |
| EPI_ISL_19048179 | GISAID | RSV-B | Canada | 2019 |
| EPI_ISL_19048188 | GISAID | RSV-B | Australia | 2019 |
| EPI_ISL_19048205 | GISAID | RSV-B | South Korea | 2019 |
| EPI_ISL_19048225 | GISAID | RSV-B | South Africa | 2019 |
| EPI_ISL_19048261 | GISAID | RSV-B | Canada | 2019 |
| EPI_ISL_19048274 | GISAID | RSV-B | Netherlands | 2019 |
| EPI_ISL_19048287 | GISAID | RSV-B | South Africa | 2019 |
| EPI_ISL_19048293 | GISAID | RSV-B | France | 2019 |
| EPI_ISL_19048307 | GISAID | RSV-B | Australia | 2018 |
| EPI_ISL_19048330 | GISAID | RSV-B | Spain | 2019 |
| EPI_ISL_19048354 | GISAID | RSV-B | South Korea | 2019 |
| EPI_ISL_19048372 | GISAID | RSV-B | Germany | 2019 |
| EPI_ISL_19048377 | GISAID | RSV-B | Netherlands | 2017 |

| | | | | |
|---|---|---|---|---|
| EPI_ISL_19048379 | GISAID | RSV-B | Australia | 2018 |
| EPI_ISL_19048381 | GISAID | RSV-B | Spain | 2019 |
| EPI_ISL_19048410 | GISAID | RSV-B | Germany | 2019 |
| EPI_ISL_19048456 | GISAID | RSV-B | Russia | 2020 |
| EPI_ISL_19048461 | GISAID | RSV-B | Russia | 2020 |
| EPI_ISL_19048479 | GISAID | RSV-B | Germany | 2019 |
| EPI_ISL_19048482 | GISAID | RSV-B | Canada | 2019 |
| EPI_ISL_19048485 | GISAID | RSV-B | Germany | 2019 |
| EPI_ISL_19048494 | GISAID | RSV-B | Spain | 2019 |
| EPI_ISL_19048511 | GISAID | RSV-B | South Africa | 2019 |
| EPI_ISL_19048512 | GISAID | RSV-B | Australia | 2019 |
| EPI_ISL_19048517 | GISAID | RSV-B | Brazil | 2018 |
| EPI_ISL_19048542 | GISAID | RSV-B | Germany | 2019 |
| EPI_ISL_19048553 | GISAID | RSV-B | South Africa | 2018 |
| EPI_ISL_19048569 | GISAID | RSV-B | United Kingdom | 2018 |
| EPI_ISL_19048573 | GISAID | RSV-B | South Korea | 2020 |
| EPI_ISL_19048583 | GISAID | RSV-B | South Africa | 2019 |
| EPI_ISL_19048587 | GISAID | RSV-B | Japan | 2019 |
| EPI_ISL_19048624 | GISAID | RSV-B | Brazil | 2019 |
| EPI_ISL_19048643 | GISAID | RSV-B | South Africa | 2018 |
| EPI_ISL_19048648 | GISAID | RSV-B | South Africa | 2019 |
| EPI_ISL_19048685 | GISAID | RSV-B | Russia | 2020 |
| EPI_ISL_19048699 | GISAID | RSV-B | Germany | 2019 |
| EPI_ISL_19048716 | GISAID | RSV-B | Brazil | 2018 |
| EPI_ISL_19048724 | GISAID | RSV-B | Germany | 2019 |
| EPI_ISL_19048734 | GISAID | RSV-B | Spain | 2018 |
| EPI_ISL_19048761 | GISAID | RSV-B | Spain | 2020 |
| EPI_ISL_19048768 | GISAID | RSV-B | South Africa | 2019 |
| EPI_ISL_19048792 | GISAID | RSV-B | Australia | 2018 |
| EPI_ISL_19048794 | GISAID | RSV-B | Canada | 2018 |
| EPI_ISL_19048817 | GISAID | RSV-B | South Africa | 2019 |
| EPI_ISL_19048861 | GISAID | RSV-B | South Africa | 2018 |

| | | | | |
|---|---|---|---|---|
| EPI_ISL_19048868 | GISAID | RSV-B | Canada | 2019 |
| EPI_ISL_19048871 | GISAID | RSV-B | Germany | 2019 |
| EPI_ISL_19048884 | GISAID | RSV-B | Germany | 2019 |
| EPI_ISL_19048912 | GISAID | RSV-B | United Kingdom | 2018 |
| EPI_ISL_19048930 | GISAID | RSV-B | Canada | 2019 |
| EPI_ISL_19048945 | GISAID | RSV-B | South Korea | 2019 |
| EPI_ISL_19048990 | GISAID | RSV-B | Brazil | 2018 |
| EPI_ISL_19049022 | GISAID | RSV-B | Spain | 2019 |
| EPI_ISL_19049047 | GISAID | RSV-B | Spain | 2018 |
| EPI_ISL_19049064 | GISAID | RSV-B | France | 2019 |
| EPI_ISL_19049070 | GISAID | RSV-B | United Kingdom | 2018 |
| EPI_ISL_19049085 | GISAID | RSV-B | South Korea | 2020 |
| EPI_ISL_19049093 | GISAID | RSV-B | South Africa | 2020 |
| EPI_ISL_19049096 | GISAID | RSV-B | Netherlands | 2017 |
| EPI_ISL_19049115 | GISAID | RSV-B | South Korea | 2019 |
| EPI_ISL_19049178 | GISAID | RSV-B | Netherlands | 2019 |
| EPI_ISL_19049219 | GISAID | RSV-B | United Kingdom | 2018 |
| EPI_ISL_19049274 | GISAID | RSV-B | United Kingdom | 2018 |
| EPI_ISL_19060412 | GISAID | RSV-B | England | 2024 |
| EPI_ISL_19063269 | GISAID | RSV-B | Italy | 2023 |
| EPI_ISL_19063275 | GISAID | RSV-B | Italy | 2019 |
| EPI_ISL_19063276 | GISAID | RSV-B | Italy | 2021 |
| EPI_ISL_19063277 | GISAID | RSV-B | Italy | 2017 |
| EPI_ISL_19063278 | GISAID | RSV-B | Italy | 2022 |
| EPI_ISL_19063279 | GISAID | RSV-B | Italy | 2017 |
| EPI_ISL_19063282 | GISAID | RSV-B | Italy | 2017 |
| EPI_ISL_19063283 | GISAID | RSV-B | Italy | 2017 |
| EPI_ISL_19063286 | GISAID | RSV-B | Italy | 2023 |
| EPI_ISL_19063289 | GISAID | RSV-B | Italy | 2019 |
| EPI_ISL_19063291 | GISAID | RSV-B | Italy | 2021 |
| EPI_ISL_19063292 | GISAID | RSV-B | Italy | 2021 |

| | | | | |
|---|---|---|---|---|
| EPI_ISL_19063294 | GISAID | RSV-B | Italy | 2019 |
| EPI_ISL_19063296 | GISAID | RSV-B | Italy | 2019 |
| EPI_ISL_19063298 | GISAID | RSV-B | Italy | 2021 |
| EPI_ISL_19063299 | GISAID | RSV-B | Italy | 2019 |
| EPI_ISL_19063302 | GISAID | RSV-B | Italy | 2018 |
| EPI_ISL_19063303 | GISAID | RSV-B | Italy | 2018 |
| EPI_ISL_19063304 | GISAID | RSV-B | Italy | 2019 |
| EPI_ISL_19063307 | GISAID | RSV-B | Italy | 2020 |
| EPI_ISL_19063308 | GISAID | RSV-B | Italy | 2019 |
| EPI_ISL_19063310 | GISAID | RSV-B | Italy | 2018 |
| EPI_ISL_19063313 | GISAID | RSV-B | Italy | 2021 |
| EPI_ISL_19063314 | GISAID | RSV-B | Italy | 2019 |
| EPI_ISL_19063316 | GISAID | RSV-B | Italy | 2021 |
| EPI_ISL_19063318 | GISAID | RSV-B | Italy | 2021 |
| EPI_ISL_19063319 | GISAID | RSV-B | Italy | 2018 |
| EPI_ISL_19063321 | GISAID | RSV-B | Italy | 2021 |
| EPI_ISL_19063323 | GISAID | RSV-B | Italy | 2021 |
| EPI_ISL_19063324 | GISAID | RSV-B | Italy | 2021 |
| EPI_ISL_19063326 | GISAID | RSV-B | Italy | 2019 |
| EPI_ISL_19063328 | GISAID | RSV-B | Italy | 2019 |
| EPI_ISL_19063329 | GISAID | RSV-B | Italy | 2021 |
| EPI_ISL_19063331 | GISAID | RSV-B | Italy | 2021 |
| EPI_ISL_19063335 | GISAID | RSV-B | Italy | 2021 |
| EPI_ISL_19063339 | GISAID | RSV-B | Italy | 2022 |
| EPI_ISL_19063344 | GISAID | RSV-B | Italy | 2021 |
| EPI_ISL_19063345 | GISAID | RSV-B | Italy | 2023 |
| EPI_ISL_19063346 | GISAID | RSV-B | Italy | 2020 |
| EPI_ISL_19063347 | GISAID | RSV-B | Italy | 2021 |
| EPI_ISL_19074734 | GISAID | RSV-B | Croatia | 2024 |
| EPI_ISL_19090940 | GISAID | RSV-B | South Korea | 2019 |
| EPI_ISL_19090962 | GISAID | RSV-B | South Korea | 2023 |
| EPI_ISL_19090963 | GISAID | RSV-B | South Korea | 2023 |

| | | | | |
|---|---|---|---|---|
| EPI_ISL_19090964 | GISAID | RSV-B | South Korea | 2023 |
| EPI_ISL_19090966 | GISAID | RSV-B | South Korea | 2023 |
| EPI_ISL_19090968 | GISAID | RSV-B | South Korea | 2023 |
| EPI_ISL_19093416 | GISAID | RSV-B | Mongolia | 2023 |
| EPI_ISL_19093422 | GISAID | RSV-B | Mongolia | 2023 |
| EPI_ISL_19125708 | GISAID | RSV-B | USA | 2020 |
| EPI_ISL_19125712 | GISAID | RSV-B | USA | 2019 |
| EPI_ISL_19125716 | GISAID | RSV-B | USA | 2022 |
| EPI_ISL_19125720 | GISAID | RSV-B | USA | 2021 |
| EPI_ISL_19125721 | GISAID | RSV-B | USA | 2020 |
| EPI_ISL_19125725 | GISAID | RSV-B | USA | 2020 |
| EPI_ISL_19125729 | GISAID | RSV-B | USA | 2021 |
| EPI_ISL_19125732 | GISAID | RSV-B | USA | 2019 |
| EPI_ISL_19125734 | GISAID | RSV-B | USA | 2019 |
| EPI_ISL_19125738 | GISAID | RSV-B | USA | 2019 |
| EPI_ISL_19125741 | GISAID | RSV-B | USA | 2021 |
| EPI_ISL_19125744 | GISAID | RSV-B | USA | 2021 |
| EPI_ISL_19125748 | GISAID | RSV-B | USA | 2021 |
| EPI_ISL_19125752 | GISAID | RSV-B | USA | 2021 |
| EPI_ISL_19125757 | GISAID | RSV-B | USA | 2020 |
| EPI_ISL_19125758 | GISAID | RSV-B | USA | 2019 |
| EPI_ISL_19125762 | GISAID | RSV-B | USA | 2021 |
| EPI_ISL_19125767 | GISAID | RSV-B | USA | 2022 |
| EPI_ISL_19125768 | GISAID | RSV-B | USA | 2019 |
| EPI_ISL_19125770 | GISAID | RSV-B | USA | 2021 |
| EPI_ISL_19125776 | GISAID | RSV-B | USA | 2019 |
| EPI_ISL_19125777 | GISAID | RSV-B | USA | 2021 |
| EPI_ISL_19125778 | GISAID | RSV-B | USA | 2021 |
| EPI_ISL_19125788 | GISAID | RSV-B | USA | 2021 |
| EPI_ISL_19125792 | GISAID | RSV-B | USA | 2021 |
| EPI_ISL_19125803 | GISAID | RSV-B | USA | 2019 |
| EPI_ISL_19125826 | GISAID | RSV-B | USA | 2021 |

| | | | | |
|---|---|---|---|---|
| EPI_ISL_19125830 | GISAID | RSV-B | USA | 2021 |
| EPI_ISL_19125831 | GISAID | RSV-B | USA | 2020 |
| EPI_ISL_19125833 | GISAID | RSV-B | USA | 2019 |
| EPI_ISL_19125835 | GISAID | RSV-B | USA | 2018 |
| EPI_ISL_19125844 | GISAID | RSV-B | USA | 2019 |
| EPI_ISL_19125845 | GISAID | RSV-B | USA | 2021 |
| EPI_ISL_19125852 | GISAID | RSV-B | USA | 2019 |
| EPI_ISL_19147781 | GISAID | RSV-B | Japan | 2023 |
| EPI_ISL_19148605 | GISAID | RSV-B | USA | 2024 |
| EPI_ISL_19159491 | GISAID | RSV-B | USA | 2024 |
| EPI_ISL_19159531 | GISAID | RSV-B | USA | 2023 |
| EPI_ISL_19160827 | GISAID | RSV-B | Ireland | 2023 |
| EPI_ISL_19160828 | GISAID | RSV-B | Ireland | 2023 |
| EPI_ISL_19160834 | GISAID | RSV-B | Ireland | 2023 |
| EPI_ISL_19206362 | GISAID | RSV-B | New Zealand | 2017 |
| EPI_ISL_19206423 | GISAID | RSV-B | New Zealand | 2017 |
| EPI_ISL_19206455 | GISAID | RSV-B | New Zealand | 2017 |
| EPI_ISL_19206457 | GISAID | RSV-B | New Zealand | 2017 |
| EPI_ISL_19206458 | GISAID | RSV-B | New Zealand | 2017 |
| EPI_ISL_19206500 | GISAID | RSV-B | New Zealand | 2017 |
| EPI_ISL_19206504 | GISAID | RSV-B | New Zealand | 2017 |
| EPI_ISL_19206506 | GISAID | RSV-B | New Zealand | 2017 |
| EPI_ISL_19206519 | GISAID | RSV-B | New Zealand | 2017 |
| EPI_ISL_19206531 | GISAID | RSV-B | New Zealand | 2017 |
| EPI_ISL_19206568 | GISAID | RSV-B | New Zealand | 2018 |
| EPI_ISL_19206572 | GISAID | RSV-B | New Zealand | 2018 |
| EPI_ISL_19206586 | GISAID | RSV-B | New Zealand | 2019 |
| EPI_ISL_19206603 | GISAID | RSV-B | New Zealand | 2018 |
| EPI_ISL_19206955 | GISAID | RSV-B | New Zealand | 2017 |
| EPI_ISL_19207016 | GISAID | RSV-B | New Zealand | 2017 |
| EPI_ISL_19207048 | GISAID | RSV-B | New Zealand | 2017 |
| EPI_ISL_19207050 | GISAID | RSV-B | New Zealand | 2017 |

| | | | | |
|---|---|---|---|---|
| EPI_ISL_19207051 | GISAID | RSV-B | New Zealand | 2017 |
| EPI_ISL_19207093 | GISAID | RSV-B | New Zealand | 2017 |
| EPI_ISL_19207097 | GISAID | RSV-B | New Zealand | 2017 |
| EPI_ISL_19207099 | GISAID | RSV-B | New Zealand | 2017 |
| EPI_ISL_19207112 | GISAID | RSV-B | New Zealand | 2017 |
| EPI_ISL_19207124 | GISAID | RSV-B | New Zealand | 2017 |
| EPI_ISL_19207161 | GISAID | RSV-B | New Zealand | 2018 |
| EPI_ISL_19207165 | GISAID | RSV-B | New Zealand | 2018 |
| EPI_ISL_19207179 | GISAID | RSV-B | New Zealand | 2019 |
| EPI_ISL_19207196 | GISAID | RSV-B | New Zealand | 2018 |
| EPI_ISL_19220729 | GISAID | RSV-B | South Africa | 2023 |
| EPI_ISL_19220730 | GISAID | RSV-B | South Africa | 2023 |
| EPI_ISL_19220734 | GISAID | RSV-B | South Africa | 2023 |
| EPI_ISL_19220737 | GISAID | RSV-B | South Africa | 2023 |
| EPI_ISL_19220740 | GISAID | RSV-B | South Africa | 2023 |
| EPI_ISL_19220751 | GISAID | RSV-B | South Africa | 2024 |
| EPI_ISL_19220752 | GISAID | RSV-B | South Africa | 2024 |
| EPI_ISL_19220753 | GISAID | RSV-B | South Africa | 2024 |
| EPI_ISL_19220755 | GISAID | RSV-B | South Africa | 2024 |
| EPI_ISL_19220756 | GISAID | RSV-B | South Africa | 2024 |
| EPI_ISL_19220757 | GISAID | RSV-B | South Africa | 2024 |
| EPI_ISL_19220763 | GISAID | RSV-B | South Africa | 2024 |
| EPI_ISL_19220765 | GISAID | RSV-B | South Africa | 2024 |
| EPI_ISL_19220769 | GISAID | RSV-B | South Africa | 2024 |
| EPI_ISL_19220792 | GISAID | RSV-B | South Africa | 2024 |
| EPI_ISL_19220793 | GISAID | RSV-B | South Africa | 2024 |
| EPI_ISL_19220794 | GISAID | RSV-B | South Africa | 2024 |
| EPI_ISL_19220810 | GISAID | RSV-B | South Africa | 2024 |
| EPI_ISL_19220819 | GISAID | RSV-B | South Africa | 2024 |
| EPI_ISL_19220824 | GISAID | RSV-B | South Africa | 2023 |
| EPI_ISL_19220826 | GISAID | RSV-B | South Africa | 2024 |
| EPI_ISL_19220827 | GISAID | RSV-B | South Africa | 2024 |

| | | | | |
|---|---|---|---|---|
| EPI_ISL_19220837 | GISAID | RSV-B | South Africa | 2023 |
| EPI_ISL_19220871 | GISAID | RSV-B | South Africa | 2024 |
| EPI_ISL_19220875 | GISAID | RSV-B | South Africa | 2024 |
| EPI_ISL_19220879 | GISAID | RSV-B | South Africa | 2024 |
| EPI_ISL_19220884 | GISAID | RSV-B | South Africa | 2024 |
| EPI_ISL_19220889 | GISAID | RSV-B | South Africa | 2024 |
| EPI_ISL_19220890 | GISAID | RSV-B | South Africa | 2024 |
| EPI_ISL_19232762 | GISAID | RSV-B | England | 2024 |
| EPI_ISL_19232763 | GISAID | RSV-B | England | 2024 |
| EPI_ISL_19232764 | GISAID | RSV-B | England | 2024 |
| EPI_ISL_19237920 | GISAID | RSV-B | USA | 2024 |
| EPI_ISL_19237922 | GISAID | RSV-B | USA | 2024 |
| EPI_ISL_19291604 | GISAID | RSV-B | England | 2022 |
| EPI_ISL_19291608 | GISAID | RSV-B | England | 2022 |
| EPI_ISL_19291611 | GISAID | RSV-B | England | 2022 |
| EPI_ISL_19291621 | GISAID | RSV-B | England | 2022 |
| EPI_ISL_19291627 | GISAID | RSV-B | England | 2022 |
| EPI_ISL_19291634 | GISAID | RSV-B | England | 2022 |
| EPI_ISL_19291637 | GISAID | RSV-B | England | 2022 |
| EPI_ISL_19291650 | GISAID | RSV-B | England | 2022 |
| EPI_ISL_19291651 | GISAID | RSV-B | England | 2022 |
| EPI_ISL_19291655 | GISAID | RSV-B | England | 2022 |
| EPI_ISL_19291657 | GISAID | RSV-B | England | 2022 |
| EPI_ISL_19291660 | GISAID | RSV-B | England | 2022 |
| EPI_ISL_19291670 | GISAID | RSV-B | England | 2022 |
| EPI_ISL_19291672 | GISAID | RSV-B | England | 2022 |
| EPI_ISL_19294764 | GISAID | RSV-B | USA | 2024 |
| EPI_ISL_19302863 | GISAID | RSV-B | England | 2024 |
| EPI_ISL_19302871 | GISAID | RSV-B | England | 2022 |
| EPI_ISL_19302873 | GISAID | RSV-B | England | 2022 |
| EPI_ISL_19302874 | GISAID | RSV-B | England | 2022 |
| EPI_ISL_19302883 | GISAID | RSV-B | England | 2022 |

| | | | | |
|---|---|---|---|---|
| EPI_ISL_19302889 | GISAID | RSV-B | England | 2023 |
| EPI_ISL_19321315 | GISAID | RSV-B | USA | 2023 |
| EPI_ISL_19321320 | GISAID | RSV-B | USA | 2024 |
| EPI_ISL_19321323 | GISAID | RSV-B | USA | 2024 |
| EPI_ISL_19321326 | GISAID | RSV-B | USA | 2024 |
| EPI_ISL_19345191 | GISAID | RSV-B | Argentina | 2024 |
| EPI_ISL_19423091 | GISAID | RSV-B | Panama | 2018 |
| EPI_ISL_19423098 | GISAID | RSV-B | Panama | 2018 |
| EPI_ISL_19442394 | GISAID | RSV-B | USA | 2021 |
| EPI_ISL_19442410 | GISAID | RSV-B | Scotland | 2022 |
| EPI_ISL_19442411 | GISAID | RSV-B | Scotland | 2022 |
| EPI_ISL_19442538 | GISAID | RSV-B | Scotland | 2020 |
| EPI_ISL_19442633 | GISAID | RSV-B | Scotland | 2021 |
| EPI_ISL_19442671 | GISAID | RSV-B | Scotland | 2022 |
| EPI_ISL_19442721 | GISAID | RSV-B | Scotland | 2021 |
| EPI_ISL_19446319 | GISAID | RSV-B | England | 2023 |
| EPI_ISL_19446320 | GISAID | RSV-B | England | 2023 |
| EPI_ISL_19446347 | GISAID | RSV-B | England | 2023 |
| EPI_ISL_19446392 | GISAID | RSV-B | England | 2024 |
| EPI_ISL_19446394 | GISAID | RSV-B | England | 2024 |
| EPI_ISL_19446395 | GISAID | RSV-B | England | 2024 |
| EPI_ISL_19446431 | GISAID | RSV-B | England | 2024 |
| EPI_ISL_19446432 | GISAID | RSV-B | England | 2024 |
| EPI_ISL_19446435 | GISAID | RSV-B | England | 2024 |
| EPI_ISL_19446438 | GISAID | RSV-B | England | 2024 |
| EPI_ISL_19446451 | GISAID | RSV-B | England | 2024 |
| EPI_ISL_19446454 | GISAID | RSV-B | England | 2024 |
| EPI_ISL_19446462 | GISAID | RSV-B | England | 2023 |
| EPI_ISL_19446469 | GISAID | RSV-B | England | 2023 |
| EPI_ISL_19493443 | GISAID | RSV-B | Cote d'Ivoire | 2023 |
| EPI_ISL_19493461 | GISAID | RSV-B | Cote d'Ivoire | 2023 |
| EPI_ISL_19493463 | GISAID | RSV-B | Cote d'Ivoire | 2023 |

| | | | | |
|---|---|---|---|---|
| EPI_ISL_19493467 | GISAID | RSV-B | Cote d'Ivoire | 2023 |
| EPI_ISL_19493471 | GISAID | RSV-B | Cote d'Ivoire | 2023 |
| EPI_ISL_19493474 | GISAID | RSV-B | Cote d'Ivoire | 2023 |
| EPI_ISL_19493477 | GISAID | RSV-B | Cote d'Ivoire | 2023 |
| EPI_ISL_19493497 | GISAID | RSV-B | Cote d'Ivoire | 2023 |
| EPI_ISL_19493498 | GISAID | RSV-B | Cote d'Ivoire | 2023 |
| EPI_ISL_19493512 | GISAID | RSV-B | Cote d'Ivoire | 2023 |
| EPI_ISL_19493519 | GISAID | RSV-B | Cote d'Ivoire | 2023 |
| EPI_ISL_19497472 | GISAID | RSV-B | Argentina | 2024 |
| EPI_ISL_19497478 | GISAID | RSV-B | Argentina | 2024 |
| EPI_ISL_19497498 | GISAID | RSV-B | Argentina | 2024 |
| EPI_ISL_19497502 | GISAID | RSV-B | Argentina | 2024 |
| EPI_ISL_19497505 | GISAID | RSV-B | Argentina | 2024 |
| EPI_ISL_19497513 | GISAID | RSV-B | Senegal | 2024 |
| EPI_ISL_19498038 | GISAID | RSV-B | Argentina | 2024 |
| EPI_ISL_19498039 | GISAID | RSV-B | Argentina | 2024 |
| EPI_ISL_19498040 | GISAID | RSV-B | Argentina | 2024 |
| EPI_ISL_19498041 | GISAID | RSV-B | Argentina | 2024 |
| EPI_ISL_19498043 | GISAID | RSV-B | Argentina | 2024 |
| EPI_ISL_19498045 | GISAID | RSV-B | Argentina | 2024 |
| EPI_ISL_19500980 | GISAID | RSV-B | Mexico | 2022 |
| EPI_ISL_19500982 | GISAID | RSV-B | Mexico | 2023 |
| EPI_ISL_19500988 | GISAID | RSV-B | Mexico | 2023 |
| EPI_ISL_19501004 | GISAID | RSV-B | Mexico | 2023 |
| EPI_ISL_19501005 | GISAID | RSV-B | Mexico | 2023 |
| EPI_ISL_19501006 | GISAID | RSV-B | Mexico | 2024 |
| EPI_ISL_19501947 | GISAID | RSV-B | USA | 2023 |
| EPI_ISL_19510711 | GISAID | RSV-B | South Africa | 2024 |
| EPI_ISL_19510712 | GISAID | RSV-B | South Africa | 2024 |
| EPI_ISL_19510726 | GISAID | RSV-B | South Africa | 2024 |
| EPI_ISL_19510729 | GISAID | RSV-B | South Africa | 2024 |
| EPI_ISL_19510730 | GISAID | RSV-B | South Africa | 2024 |

| | | | | |
|---|---|---|---|---|
| EPI_ISL_19510747 | GISAID | RSV-B | South Africa | 2024 |
| EPI_ISL_19510768 | GISAID | RSV-B | South Africa | 2024 |
| EPI_ISL_19510782 | GISAID | RSV-B | South Africa | 2024 |
| EPI_ISL_19510788 | GISAID | RSV-B | South Africa | 2024 |
| EPI_ISL_19517139 | GISAID | RSV-B | USA | 2023 |
| EPI_ISL_19517156 | GISAID | RSV-B | USA | 2024 |
| EPI_ISL_19517162 | GISAID | RSV-B | USA | 2023 |
| EPI_ISL_19517164 | GISAID | RSV-B | USA | 2023 |
| EPI_ISL_19517168 | GISAID | RSV-B | USA | 2023 |
| EPI_ISL_2558987 | GISAID | RSV-B | Switzerland | 2019 |
| EPI_ISL_2575511 | GISAID | RSV-B | Australia | 2016 |
| EPI_ISL_2588785 | GISAID | RSV-B | Japan | 2017 |
| EPI_ISL_732366 | GISAID | RSV-B | England | 2018 |
| EPI_ISL_732371 | GISAID | RSV-B | England | 2018 |
| OQ024160.1 | Genbank | RSV-B | USA | 2022 |
| OQ024162.1 | Genbank | RSV-B | USA | 2022 |
| OQ171933.1 | Genbank | RSV-B | USA | 2022 |
| OQ171934.1 | Genbank | RSV-B | USA | 2022 |
| OQ171937.1 | Genbank | RSV-B | USA | 2022 |
| OQ171938.1 | Genbank | RSV-B | USA | 2022 |
| OQ171942.1 | Genbank | RSV-B | USA | 2022 |
